# Understanding community level influences on the prevalence of SARS-CoV-2 infection in England

**DOI:** 10.1101/2022.04.14.22273759

**Authors:** Chaitanya Joshi, Arif Ali, Thomas ÓConnor, Li Chen, Kaveh Jahanshahi

## Abstract

Understanding and monitoring the major influences on SARS-CoV-2 prevalence is essential to inform policy making and devise appropriate packages of non-pharmaceutical interventions (NPIs). Through evaluating community level influences on the prevalence of SARS-CoV-2 infection and their spatiotemporal variations in England, this study aims to provide some insights into the most important risk parameters. We used spatial clusters developed in Jahanshahi and Jin, 2021 as geographical areas with distinct land use and travel patterns. We also segmented our data by time periods to control for changes in policies or development of the disease over the course of the pandemic. We then used multivariate linear regression to identify influences driving infections within the clusters and to compare the variations of those between the clusters. Our findings demonstrate the key roles that workplace and commuting modes have had on some of the sections of the working population after accounting for several interrelated influences including mobility and vaccination. We found communities of workers in care homes and warehouses and to a lesser extent textile and ready meal industries and those who rely more on public transport for commuting tend to carry a higher risk of infection across all residential area types and time periods.

## 1 Background

The response of governments across the globe to COVID-19 pandemic has included non-pharmaceutical interventions (NPIs). In the UK, those include mobility restrictions, closures of some industrial sectors and schools, social distancing and mandatory face covering in public areas and public transport. Assessing the effectiveness of these policies requires a thorough understanding of risk factors and behavioural responses which tends to vary over time, and across areas and communities. In response, this paper focuses on exploring the spatial and temporal (from 4^th^ October 2020-5^th^ December 2021) variations of SARS-CoV-2 infection risk factors at aggregate geographical level.

Over the last couple of years, many studies have focused on individual and household level influences for SARS-CoV-2 infection and associated outcomes (e.g. House et al, 2021; Williamson, E.J et al, 2020; Katikireddi et al, 2021; Jin J et al, 2021; Sze S et al, 2020). For instance, House et al (2021) used the Office for National Statistics (ONS) COVID-19 Infection Survey (CIS)^1^ to evaluate within and between household transmission and the associated factors ranging from socioeconomic and demographic characteristics to genome structures and vaccination records^2^. Williamson et al (2020) reported one of the earliest studies outlining risk factors that are associated with COVID-19-related deaths in England^3^. Katikireddi et al (2021) highlighted the interplay of individual demographic factors based around ethnicity which contributes to certain groups having unequal risk exposure to COVID-19 such as household sizes, disease vulnerability and occupation^4^. Raisi-Estabragh et al (2020) investigated the heightened risk of COVID-19 to Black and Asian ethnicities using data from the UK Biobank (UKB) highlighting the importance of risk influences such as material deprivation and housing conditions^5^. Jin et al (2021) devised a population risk calculator for COVID-19 mortality based on various sociodemographic factors and pre-existing conditions^6^. Sze S. et al (2020) reviewed existing works on the association of ethnicity with vulnerability to COVID-19 infection and clinical outcomes at the individual level^7^. Going beyond inferring COVID-19 risk factors at individual level, there have been limited investigations mapping and identifying community risk factors for COVID-19 (Khunti 2020, Wang 2021)^8,9^. Examples are Khunti et al (2020) which pointed out potential association between ethnicity and outcome in COVID-19 in ethnically diverse communities such as in the UK^8^ and Wang (2021) which explored the risk factors for nursing home COVID-19 death rates^9^. Whilst person level (dis-aggregate) analyses mainly consider individual and household characteristics, the community level analysis focuses on the make-up of an area and can account for interactions between socioeconomic profile, mobility patterns, and land use features of neighbourhood and settlements. Consequently, this can offer an alternative perspective for comprehending vulnerable regions and communities, enabling the implementation of more focused policies and direct follow-up dis-aggregate analysis. For example, analysis of the ONS individual level COVID-19 risk screening model^10^ helps identify characteristics of people who are more likely to test positive for COVID-19 in specific periods of time (e.g. those who live in four person household compared to those living alone). On the other hand, a community level risk analysis could potentially look at how the proportion of multi-person households within a Lower Layer Super Output Area (LSOA)^1^ affects the risk of infection after accounting for other area specific features such as vaccination rate, mobility patterns or land use characteristics. To the best of our knowledge, we are not aware of any prior investigation which comprehensively evaluates community level risks of SARS-CoV-2 infection after controlling for areas’ associated characteristics (including real time mobility patterns); this forms the motivation of our present investigation.

## 2 Objectives

This study aims to evaluate community level (area based) associated risks to the prevalence of SARS-CoV-2 infection. Specifically, we aim to understand the relative risks of areas with higher proportion of those working in high risk industrial sectors after controlling for other area specific risk factors such as the land use characteristics, vaccination rate, and mobility patterns. Pandemic tends to behave differently across communities; for instance, the rate and number of infections is evidenced to be higher in dense urbanised areas when compared to more rural neighbourhoods or among certain ethnic groups such as Asians. However, spatial interactions and interrelated influences makes it difficult to drive causal inference. As an example, the residential preferences or spatial sorting of Asians might mean that they tend to live in high dense and urbanised areas makes it difficult to conclude if it is ethnicity or rather area type or higher population density which results in higher risk of infection for Asians. In the context of evaluating workplace risks, it is of policy interest to understand whether the greater risk of infection for certain high risk industries, such as warehouse workers, is due to the areas where those workers are residing, factors related to the ethnicity of workers in those jobs, or the settings of the workplace. Understanding the most important factors after controlling for a wide range of community level influences and accounting for spatial interactions and temporal variations is the main motivation behind this study.

## 3 Methods

A broad range of area based influences can contribute to SARS-CoV-2 infection including socioeconomic and demographic profile, built-form characteristics, and trends in mobility and vaccination rates. Also, as stated in previous sections, these influences tend to be highly interrelated making causal inference more difficult. Ideally, we require a dataset that not only cover an extensive range of the influences at community level, but also provide a structured system-level understanding. This is a very tall order indeed, and the requirements are unlikely to be satisfied by a single data source.

To address this challenge, we assembled a variety of static (socioeconomic and demographic profile and land use characteristics) and dynamic (mobility indicators, SARS-CoV-2 positive tests and vaccination uptake in real time) features at high geographical resolution from a range of data sources in England. Methodologically, we combined machine learning and statistical techniques to a) segment our data by distinct area types, travel patterns and time periods and account for interrelated influences, and b) analyse and compare influences on the prevalence of SARS-CoV-2 infection (the outcome variable) across those more homogeneous and distinct clusters.

### 3.1 Data sources

We assembled and combined the required data from a wide range of data sources including Census 2011, 2019 Mid-Year population estimates, Inter-Department Business Register (IDBR) 2019 dataset on workplace and industrial sectors, the national travel survey (NTS) 2002 to 2015 to segment travel patterns and clusters, mobile phone data to extract real-time mobility indicators, uptake of first and second doses of COVID-19 vaccination and test and trace data for gathering dynamic information on the prevalence of SARS-CoV-2 infection. The list of the variables is arguably the most comprehensive in England for LSOA level analysis of the prevalence of SARS-CoV-2.

Figure 1 shows a schematic of datasets used in analysis of community level influences. The datasets can be categorised into static, which are assumed to be constant over time, and dynamic, which do change over time. Further details on the list of variables and the break down of levels are provided in Tables A.2 in Appendix A. A schematic of data linking process is shown in Figure 2.

**Figure 1.**
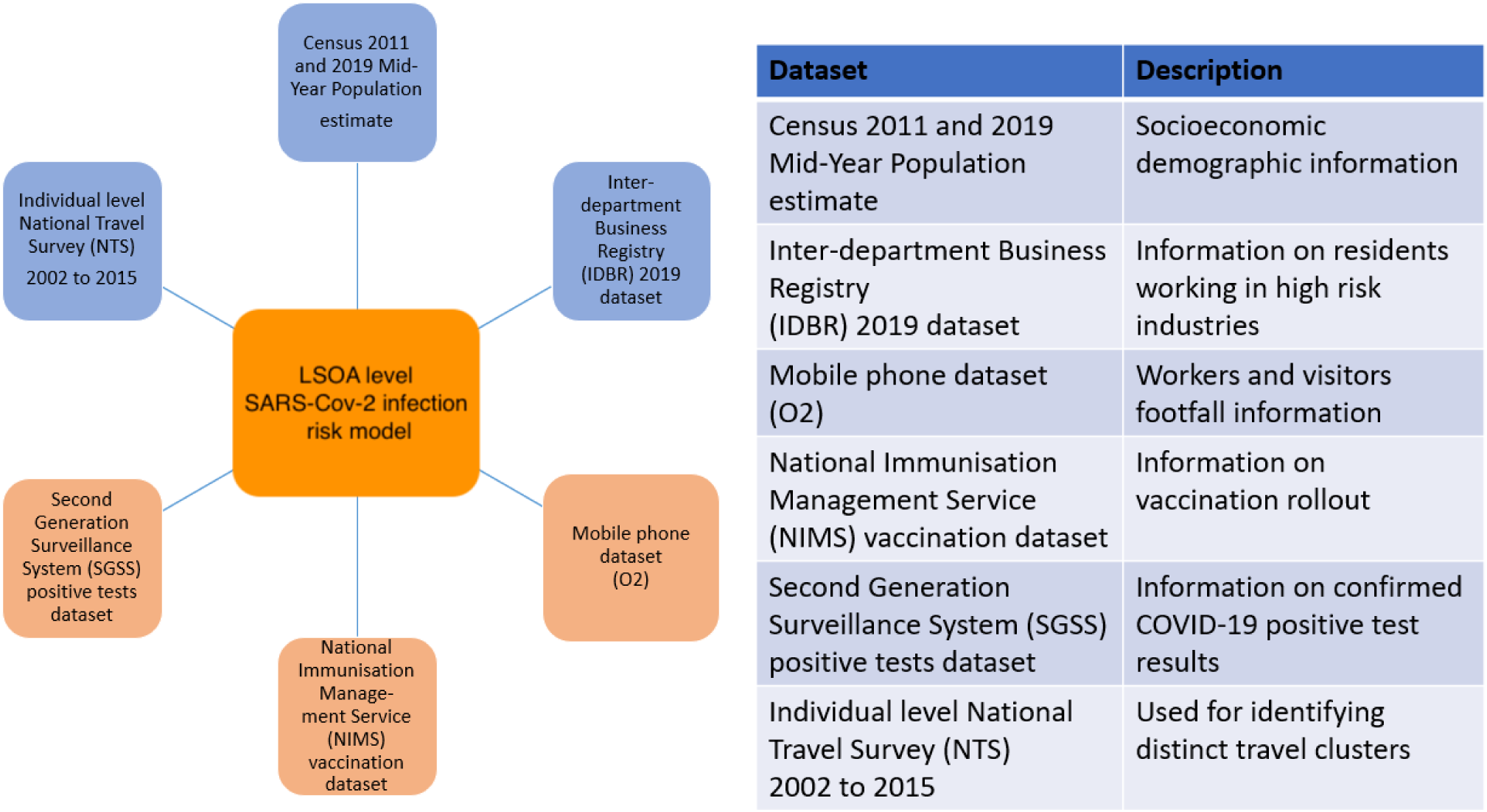
The main data sources used in the analysis alongside their respective descriptions. The spatial and temporal resolutions of the data are presented in Table A.3 in Appendix A.

**Figure 2.**
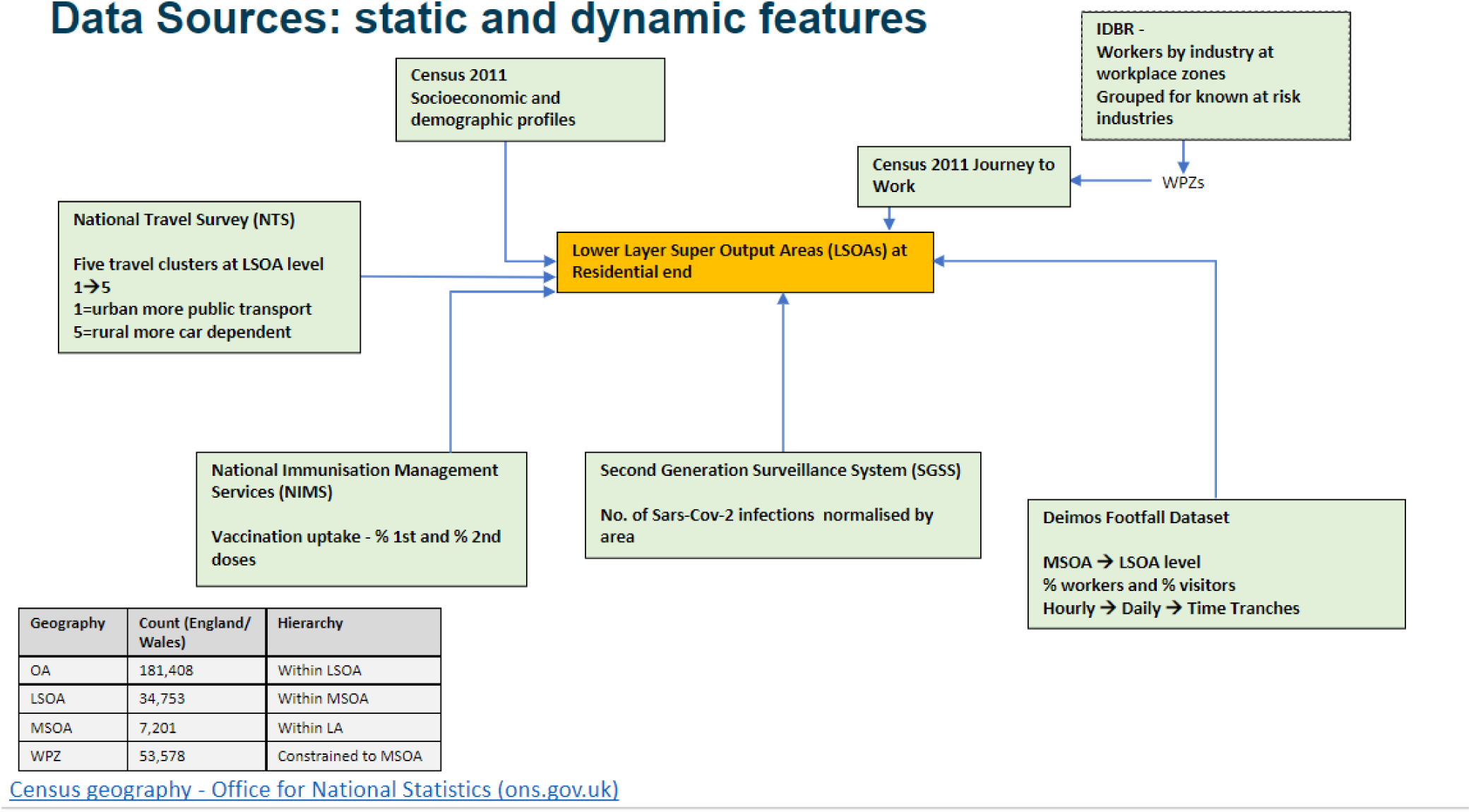
The data linking process linking the various static and dynamic data sources along with different UK Census geographies.

#### 3.1.1 Static exposure variables

The main static variables were derived from Census 2011^12–14^, which albeit being 10 years old, still provides a rich source of data and detailed snapshot on socioeconomic and demographic profiles of LSOAs ^2^. Nevertheless, where possible, data was supplanted by more recent sources such as 2019 population estimates. More detailed description of each variable used in our model is provided in Table A.2.

We also used the Inter-Departmental Business Registry (IDBR) 2019^15^ to extract information on certain industry types namely meat and fish processing, textiles, care homes ^3^, warehousing and ready meals. These group of industries were identified as industries of potential interest associated with some of the highest SARS-CoV-2 infection rates ^4^ in the early stages of the pandemic in the UK (we will refer to them as high-risk industries for the rest of this paper)^16^. The analysis, however, can be expanded to include other industrial sectors. The IDBR is a comprehensive register of UK businesses used by government for statistical purposes and provides the main sampling frame for surveys of businesses carried out by the ONS and other government departments. The two main sources of input for IDBR are Value Added Tax (VAT) and Pay As You Earn (PAYE) records from HMRC. Additional information comes from Companies House, Dun and Bradstreet and ONS business surveys. The IDBR covers around 2.7 million businesses in all sectors of the economy, but since the main two tax sources have thresholds, very small businesses operating below these will, in most cases, not be included. This is unlikely to be a major problem for our analysis as the high-risk industries we have modelled are unlikely to include many of the small size businesses below the tax threshold. The IDBR 2019 data was aggregated to a workplace zone (WPZ) geography^17^ and each row in the aggregated table provides the Standard Industrial Classification (SIC) code and name, industrial classification group code, WPZ, and the number of individual employed at the corresponding SIC^18^. Table A.4 in Appendix A lists the SIC codes of the high-risk industries used in this paper.

The Census 2011 Journey to work dataset provides the location of usual residents living at Output Area and working at WPZ^5^. We merged the Census 2011 travel-to-work dataset and IDBR 2019 dataset to produce the number of residents living at the Output Area level by industrial sectors in which they work. Output Area data is then aggregated to LSOA to create LSOA level of working population by industry type needed for our analysis. Finally, for the purpose of modelling, the number of high-risk industries’ workers was normalised by LSOA area and expressed in area adjusted units (per Hectare).

#### 3.1.2 Dynamic exposure variables

One of the novel aspects of our analysis was to incorporate dynamic mobility and vaccination data and augment that with static socioeconomic and demographic and built-form features. For this analysis, we used work and visiting footfall data from Telefonica (we call this Deimos data). Deimos footfall provides information on the number of devices counted at the Middle Layer Super Output Areas (MSOA) overlayed with behavioural insights broken by different age bands, gender, and travel purposes (visitor, worker and resident) on an hourly basis. The data that we had access to was anonymised and aggregated, never allowed for identification or mapping of individuals and no personal information could be identified. We aggregated the data from hourly to daily counts and further average for each modelled time tranche (see Table A.5), and then dis-aggregated the counts from MSOA to LSOA geographical level, using the total number of workers at workplace (from IDBR data) normalised by area as weights.

The second dynamic dataset was the vaccination data reported by the National Immunisation Management Service (NIMS). The dataset contained anonymised individual level information including the vaccine dose the person received (their first or second), the date, and their age. For our model, we considered the total number of administrated doses at the end of each successive time tranche for each individual LSOA normalised by the population of each individual LSOA (using 2019 population Mid-year estimate). There are some known issues with NIMS data specifically when one wishes to aggregate and estimate the proportion of those vaccinated. NIMS may over-estimate denominators in some age groups, for example because people are registered with the NHS but may have moved overseas^19^. Using 2019 population Mid-year estimate can also erroneously lead to greater than 100% vaccine coverage for some age groups. To minimise some of the biases, we focused on the rate of administration of second dose after the first dose (the difference between first and second doses within given time tranches). We constructed a single variable to collectively account for the number of first and second vaccination doses administered within each time tranche and each LSOA defined as follows:

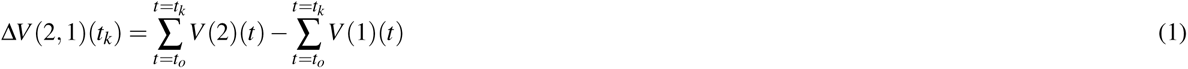

where *V* (1)(*t*) and *V* (2)(*t*) are LSOA aggregated cumulative proportion of population administered with the first and second doses of COVID-19 vaccination correspondingly within each time tranche, *t* and Δ*V* (2, 1)(*t_k_*) represents the difference between the proportion of the fully vaccinated and partially vaccinated population for each successive time tranche (refer to Table A.5 for the definition of time tranches). In addition to the better chance of cancelling out the biases through incorporating the difference in the cumulative administrated doses, the constructed variable (rate of being fully vaccinated) is most relevant to controlling prevalence of SARS-CoV-2 infection, our target variable. Other investigations have suggested that one dose of vaccine cannot fully immunise people against the infection and that vaccination protection against the risk of catching SARS-CoV-2 infection slides over time^20,21^.

#### 3.1.3 Dynamic outcome variable

Finally, the dataset on confirmed COVID-19 positive test results was provided to us by Public Health England (PHE) and include the Second Generation Surveillance System (SGSS). SGSS contains all positive specimens for any notifiable disease in England, including all pillar 2^6^ positives which includes wider population tests outside of NHS labs and hospitals such as through regional test sites, home testing kits and mobile testing sites^7^. These are subsequently transformed into case records as appropriate for a given disease. This dataset holds records on an individual level of positive test results while providing demographic information of the individuals such as age and gender. Individuals are anonymised, however, geographic information of the individual’s residence is provided at a LSOA level. For the purposes of this research, this dataset was aggregated to a LSOA geography and normalised by area (expressed as per square km) for each individual LSOA; this data served as the dependent variable for our model. It is worth highlighting that although LSOA aggregated COVID-19 positive test results can be one of the most reliable indicator of the state of the epidemic, the dataset has its own limitations^22^. For instance, it has been shown that symptomatic testing is likely to underrepresent younger population. Other testing biases reported in the literature include accessibility, reporting lags, and the ethical aspect upon receiving a positive result.

### 3.2 Statistical methods

Figure 3 shows our LSOA community risk modelling framework. We adopted a two stage process: latent cluster analysis (LCA) to capture distinct travel and land use clusters across the country (left hand side graph in Figure 3), and a multigroup multivariate regression to estimate influences on the prevalence of SARS-CoV-2 infection within each identified travel cluster (the right hand side graph in Figure 3). We also employed Exploratory Factor Analysis (EFA) to explore and combine the interrelated variables to construct new input variables where necessary. This is further explained in Section B.1.

**Figure 3.**
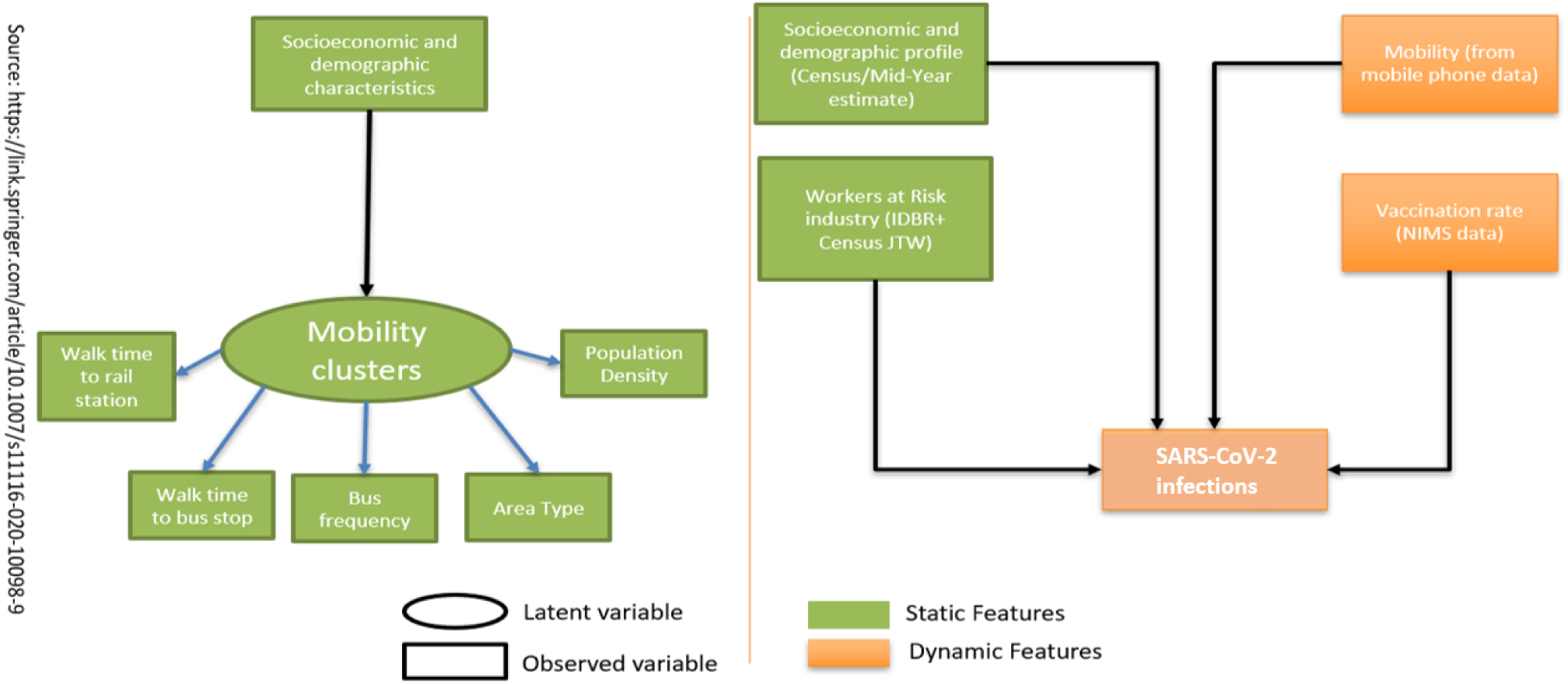
Structure of the underlying LSOA community risk model. (Left) Conditional Latent Cluster Model (based on National Travel Survey data). (Right) Multi-variate regression model for each latent cluster and time tranche.

The LCA is based on our earlier work^11^ where we analysed a wide range of built form indicators (namely area type, population density, walk time to bus stop and rail station, and bus frequency) and socioeconomic characteristics from the individual level National Travel Survey (NTS) data and identified five distinct travel clusters within which exist more homogenous land use patterns and travel attitudes. Based on their characteristics, we labelled the clusters as: L1 - Metropolitan core dwellers (mainly Inner and Central London), L2 - outer metropolitan dwellers (mainly outer London and Metropolitan areas), L3 - suburban dwellers, L4-exurban dwellers and L5-rural dwellers (Figure B.1(a) presents the geographical distribution of travel clusters).

We developed separate models for each travel cluster (multigroup modelling approach) to account for heterogeneities across geographical areas. First, through multigroup approach, we accounted for potential self-selection and spatial sorting effects reflected in the tendency of socioeconomic and demographic groups to reside in residential areas based on their land use, travel preferences, social structure, or social inequalities (e.g. as shown in Section B.1, Asian/ Asian British ethnicity group has a higher tendency to live in metropolitan cities). Through segmenting by travel clusters, we accounted for the effect of residential areas’ built-form characteristics (e.g. area type and associated travel patterns) when analysing the impact of other influences (e.g. proportion of Asians and Asian British) on the risk of SARS-CoV-2 infection. Second, different travel clusters have potentially very different associated risks of infection; for instance, the SARS-CoV-2 infection risk factors in dense urbanised areas with larger levels of mobility and more diverse socioeconomic and demographic profile of residents (e.g. London) are likely to be very different from those in small/medium urban or rural areas.

#### 3.2.1 Conditional Latent Cluster Analysis

Conditional LCA involves allocating individuals to distinct clusters in the way that it maximises similarity within clusters and the differences between clusters based on individuals’ residential built environment characteristics and conditional on their socioeconomic and demographic profile.

To formulate, let, 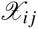 be the *j^th^* indicator variable (e.g.. population density, area type, etc) of the travel cluster, *C_i_* for individual *i*. As all our indicators are ordered categorical variables, we can formulate the link function by defining an underlying continuous variable, 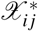 such that

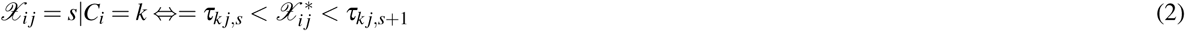

where *s* is one of the possible categorical values for, 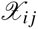, *C_i_*, our travel cluster variable which takes a value between 1,…,*k* and *τ* are a set of threshold parameters. Conditional on regressors *X* (i.e. socioeconomic characteristics ^8^ in our case) we can then present the link function as:

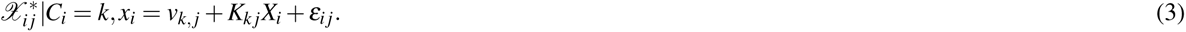

The normal distribution assumption for *ε_i j_* is equivalent to a probit regression for categorical variable *χ_i j_* on *X_i_* with the following probability function:

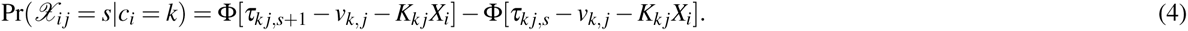

Finally, the travel cluster membership probability conditional on *X* is given by multinomial logistic regression with the following formula:

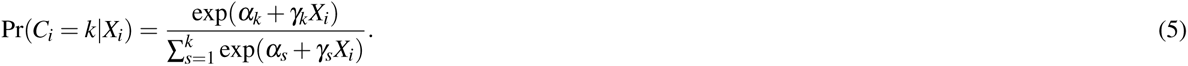

#### 3.2.2 Segmenting by time periods

In addition to segmenting by travel clusters (spatial segmentation), we also split the time series into segments to represent different policy interventions (NPI regimes) and assess the variations in influences over time^2^. This also helps account for the heterogeneity from external influences (e.g. the new variant of virus). Each time tranche corresponds to a period defined by a specific external influence and NPI and allows us to model our data more homogeneously. The distinct tranches under exploration are shown in Table A.5.

#### 3.2.3 Exploratory Factor Analysis

We employed exploratory factor analysis (EFA) on our input variables in order to explore and account for potential spatial interactions among predictors (e.g. the likelihood of workers in textile and ready meat industries to live near each other). EFA is a statistical method^23^ to reduce dimensionality of our variables through estimating the minimum number of unobserved factors that represent the observed variables. In other words, EFA describes variability among observed correlated variables in terms of a potentially lower number of unobserved variables called factors. In our case, we used EFA to evaluate the interaction between our variables and decide how to account for those as inputs to our model. To formulate, we considered each observable variable (factor indicator) *X_i_* as a linear function of independent factors and error terms which can be written as

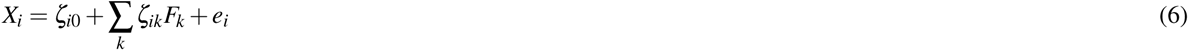

In the equation above *ζ_i_*_0_ is the bias term and the error terms *e_i_*, serve to indicate that the hypothesised relationships are not exact. In the vocabulary of factor analysis, the parameters *ζ_ik_*is referred to as factor loading of variable *X_i_* on factor *F_k_*. It can be shown that the variance of *X_i_*consists of two parts

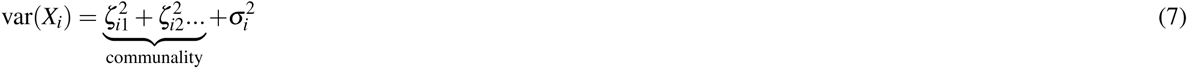

The first part, the communality of the variable, is the part that is explained by the common factors *F_k_*(*k* = 1, 2*, …N*). The second part, the specific variance, is the part of the variance of *X_i_* that is not accounted for by the common factors. One of the prime goals in exploratory factor analysis is to determine a set of loadings which bring the estimate of the total communality as close as possible to the total of the observed variances.

To perform EFAs on our dataset, we used Python’s FactorAnalyzer package^24^ and used varimax rotation on the dataset. We chose a loading threshold^25^ of 0.35 to obtain the variables contributing most to each individual factor. Factor analysis allowed us to gain invaluable insights into the dataset and create new variables to account for spatial interactions in inferring major risk factors of SARS-CoV-2 infection at the community level. Details of the results of the factor analysis are discussed in Appendix B - Section B.1.

In summary, the main assumptions made based on the findings from EFA are:

- Due to strong spatial correlation between residents working in care homes and warehouses, we combined the two as ‘total working in care home and warehouses’.
- For the same reason, we defined a new variable capturing the total resident population working in ready meals and textiles industries.

#### 3.2.4 Multigroup Multivariate linear regression at each time tranche

For each travel cluster and time tranche, we fitted a multivariate linear regression model^23^ using static and dynamic predictors as the independent variables and area adjusted SARS-CoV-2 infections as the target variable. We tested linear regression with and without regularisation and found that Linear regression approach with and without regularisation provided qualitatively similar results. Including regularisation, although it helps avoid overfitting in forecasting, produces somewhat biased estimation as it adds an extra term to the cost function. Therefore, we decided to use linear regression without regularisation to report unbiased significant risk predictors and associated p-values. Linear regression with added regularisation was used to predict area adjusted SARS-CoV-2 infections on unseen test data where unbiased estimates of the coefficients is not of primary interest^9^.

## 4 Results

In this section we are interested in uncovering the significant SARS-CoV-2 infection risk factors and their evolvement over time and evaluating the role of workplaces’ industrial sectors after controlling for a wide range of static and dynamic variables. However, before reporting the major risk influences over time and across travel clusters, we want to highlight that there exists significant spatial interaction amongst the input variables. A detailed analysis to account for these interactions is provided in Appendix B. It is worth highlighting that distribution of different predictors modelled in this study vary significantly by different clusters and these are shown in Figures B.1–B.4 in Appendix B. The analysis presented in Appendix B reveal the spatial distribution of static and dynamic predictors and show the importance of adopting a multi-group approach; analysing more homogenous data at each travel cluster better captures linear impacts when interrelations with geography is controlled. Through accounting for residential location characteristics and associated travel patterns, we can move towards making causal inference after controlling for externalities from self-selection and spatial sorting. For instance, without segmenting by travel clusters, in analysing those who work in meat and fish processing industries, we would not have been able to distinguish the impact of living in dense urbanised areas from proportion of working in meat and fish processing as those who work in this industrial sector have also higher probability of living in dense urbanised areas. Segmenting by travel clusters, on the other hand, allows separating these impacts and compare the risk of working in meat and fish processing across different built-form settings.

### 4.1 Main findings: Identifying Risk factors and their interpretation

Table 1 presents the findings from multi-group linear regression for each travel cluster and time tranche. Non-standardised coefficients, measured in their original scales, of static and dynamic influences in addition to their level of significance are reported alongside 95% confidence intervals. Non-standardised coefficients show the scale of change in a dependent variable for one unit of change in an independent variable keeping all other independent variables constant. For instance, the value of 30.82 in Table 1 shows that in the second time tranche for travel cluster 1, when all other influences remain constant, one unit increase in the proportion of Asian and Asian British ethnicity group results in 30.82 unit increase in the number of SARS-CoV-2 infections per square km. Non-standardised coefficients obtained using a multi-group approach allows us to assess relative risk of infection for different travel clusters. For example, it can be concluded from Table 1 that the relative risk of infection in areas with a large proportion of workers in high-risk industries, e.g. care homes and warehouses, is highest in most dense urbanised areas.

**Table 1.**
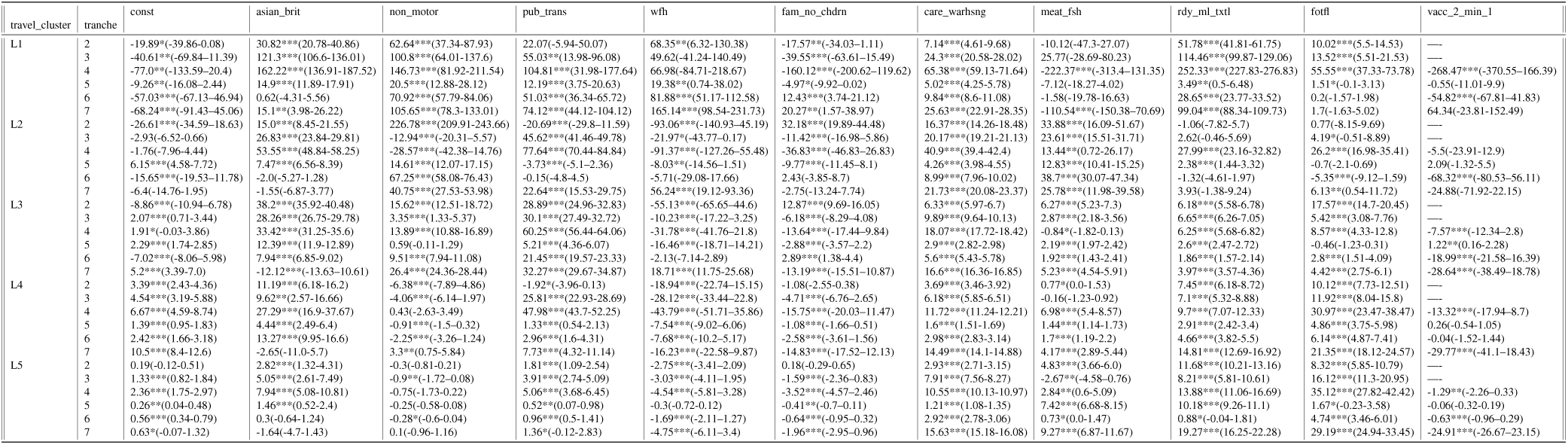
Non-Standardised risk predictors influencing infections in England covering the period 4^th^ October 2020-5^th^ December 2021 *∗∗∗p <* 0.01*, ∗∗ p <* 0.05*, ∗p <* 0.1. The predictors have been explicitly described in Table 2. Also provided are the 95% confidence interval for each coefficient. Since the vaccination coverage was negligible up until time tranche 4, we have excluded vaccination variable from the modelling for the tranches 2 and 3. Additionally, as noted in the main text the data sharing agreement with our Mobility footfall data provider does not include the period under tranche 1 and this is why tranche 1 has been excluded from the modelling.

**Table 2.**
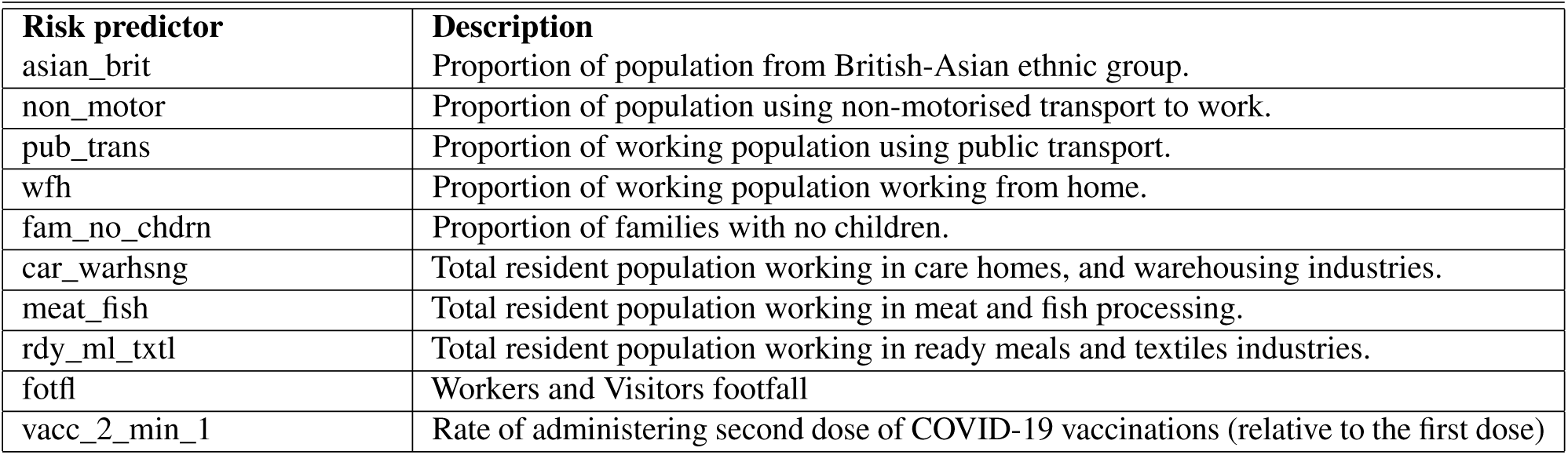
Description of the static and dynamic risk factors presented in Table 1.

However, not all variables in our model are measured on the same scale and we need to use the standardised version of their coefficients to compare different variables with each other. In terms of standardised coefficients, a change of one standard deviation in the predictor is associated with a change in the standard deviations of the dependent variable with the magnitude of the corresponding standardised coefficient. Standardised risk influences driving transmissions during the most recent time tranche (covering the period since the lifting of lockdown restrictions and before the Omicron variant became dominant in England-between the 18^th^ July 2021 and the 5^th^ December 2021) is shown in Figure 4. Standardised coefficients for other travel clusters and time tranche are presented in Table C.1. This allows us to additionally compare relative importance of different (now unitless) predictors within a given travel cluster and time tranche.

**Figure 4.**
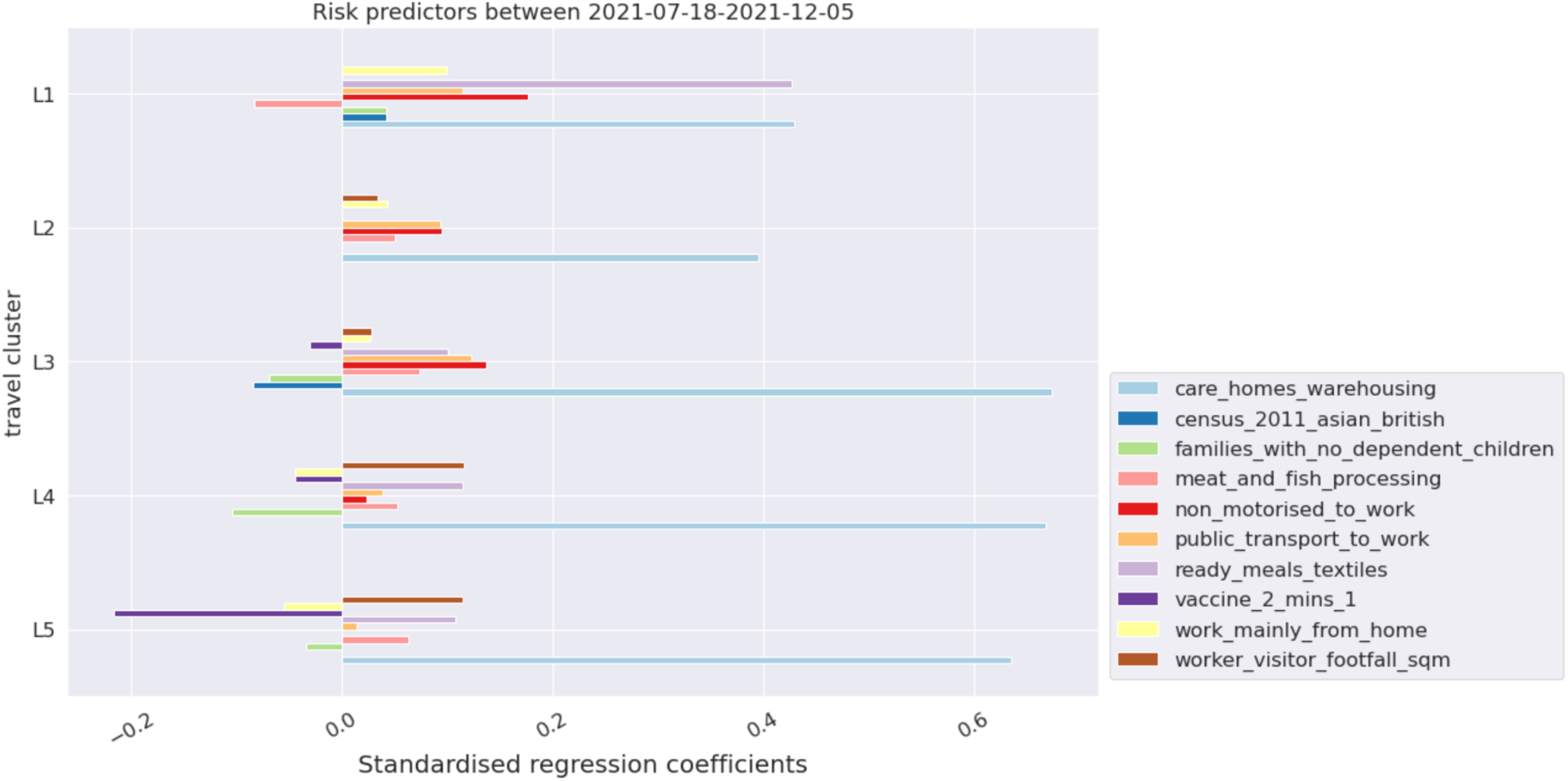
Significant risk predictors influencing infections in England for time tranche 7 corresponding to the lifting of lockdown restrictions covering the period 18^th^ July 2021-5^th^ December 2021. The coefficients are standardised and *∗p <* 0.1.

From Figure 4, it is relatively straightforward to assess risk factors for each travel cluster for the latest period reported in this paper (i.e. between 18^th^ of July 2021 to 5^th^ of December 2021). One of the key observations is that areas with a higher proportion of residents working in high-risk industries are among those most at the risk of infection. This includes both residents working in care homes and warehouses as well as those working in ready meals and textile industries. Amongst all the risk influences, areas with residents working in these industries were the dominant risk factor in both the dense urbanised and rural areas (see light blue bars in Figure 4). This is also the case across all other time tranches (refer to Table C.1 in Appendix C which shows standardised coefficients across all time tranches). The fact that the relative risk of infection in these specific industries is significant in all travel clusters suggests that some major part of the risk might stem from workplace or job requirements. In addition, we can make the following observations:

- Areas with smaller family sizes with no dependent children are less at risk of infections compared to those with a higher proportion of larger families with children. This is found to be true for suburban, exurban and rural dwellers for the most recent time tranche. An exception to this were metropolitan dwellers (inner and central London), where areas with a greater proportion of smaller family sizes with no dependent children were found to be at an increased risk of infections for the most recent time tranches (6,7) potentially associated with role of mobilities and greater mixing in London amongst its younger population.
- Areas with a higher proportion of public transport users and non-motorised commuters are more at risk. The fact that this is the case in different travel clusters (both densely and sparsely populated areas) means the risk is more likely associated with public transport and not only limited to land use features of the areas. This risk also tends to increase in most recent time tranches (as mobility increases and restrictions lifted) as can be evidenced from Appendix D.
- Areas with higher tendency to work from home are associated with lower risk of infection for more rural areas. For central and inner London (and for outer London and suburban dwellers since the lifting of lockdown restrictions), a higher proportion of the working population working from home is not significantly associated with reduced risk of infection (specifically in most recent time tranches aligned with ease in restrictions). This can be associated with a complex interplay between other purposes and forms of mobilities in more dense urbanised areas, change in human behaviour post vaccination roll-out and greater mixing following lifting of lockdown restrictions.
- As expected, in all travel clusters (land use settlements), increase in visiting and working footfall is significantly associated with higher risk of infection. This is found to be true for each individual time tranche explored in this study.
- Areas with higher rate of administering second dose of COVID-19 vaccinations (relative to the first dose of COVID-19 infection) are associated with lower risk of infection. Since the lifting of lockdown restrictions in England, this has been found to be statistically significant for the majority of population living in suburban, exurban and rural dwellers (travel clusters L3, L4, L5) which have the highest proportion of fully vaccinated populations in England by the date of this study.

### 4.2 Other analysis: Variability of Risk predictors

In the previous section, we presented the significant risk predictors for separate time tranches reflecting different policy interventions and behaviour of the pandemic. The identified risk factors stability can also be studied collectively over the entire time period. We can check the coefficient variability through *k−*fold cross-validation which is a form of data perturbation. *k−*fold divides the data into *k* non-overlapping parts (called folds), hold out one of these folds and use the remaining folds (*k −* 1) to train a model^26^. This process is repeated across all the folds until all the data has been used. To reduce potential biases, one can also repeat the *k−*fold cross-validation procedure multiple times and report the results across all folds from all runs.

Figure 5 reports the regression coefficients obtained for all folds from all runs. If estimated coefficients vary significantly when changing the input dataset their robustness is not guaranteed and they should probably be interpreted with caution. In our case, in addition to checking the robustness of results for specific time tranches (refer to Figure 5a which shows the stability of both static and dynamic influences for the latest time tranche), we also used this technique to evaluate the variations in static influences across the whole study period within each travel cluster (refer to Figure 5b^10^). The latter shows the extent to which NPIs, virus variants and other external factors might have affected the direction of impact and the relative importance of static influences. For instance, we can evaluate whether in-risk industries have had different direction of impacts on risk of infection in different time periods or whether their relative importance compared to other influences has changed over time, say due to adoption of certain policies.

**Figure 5.**
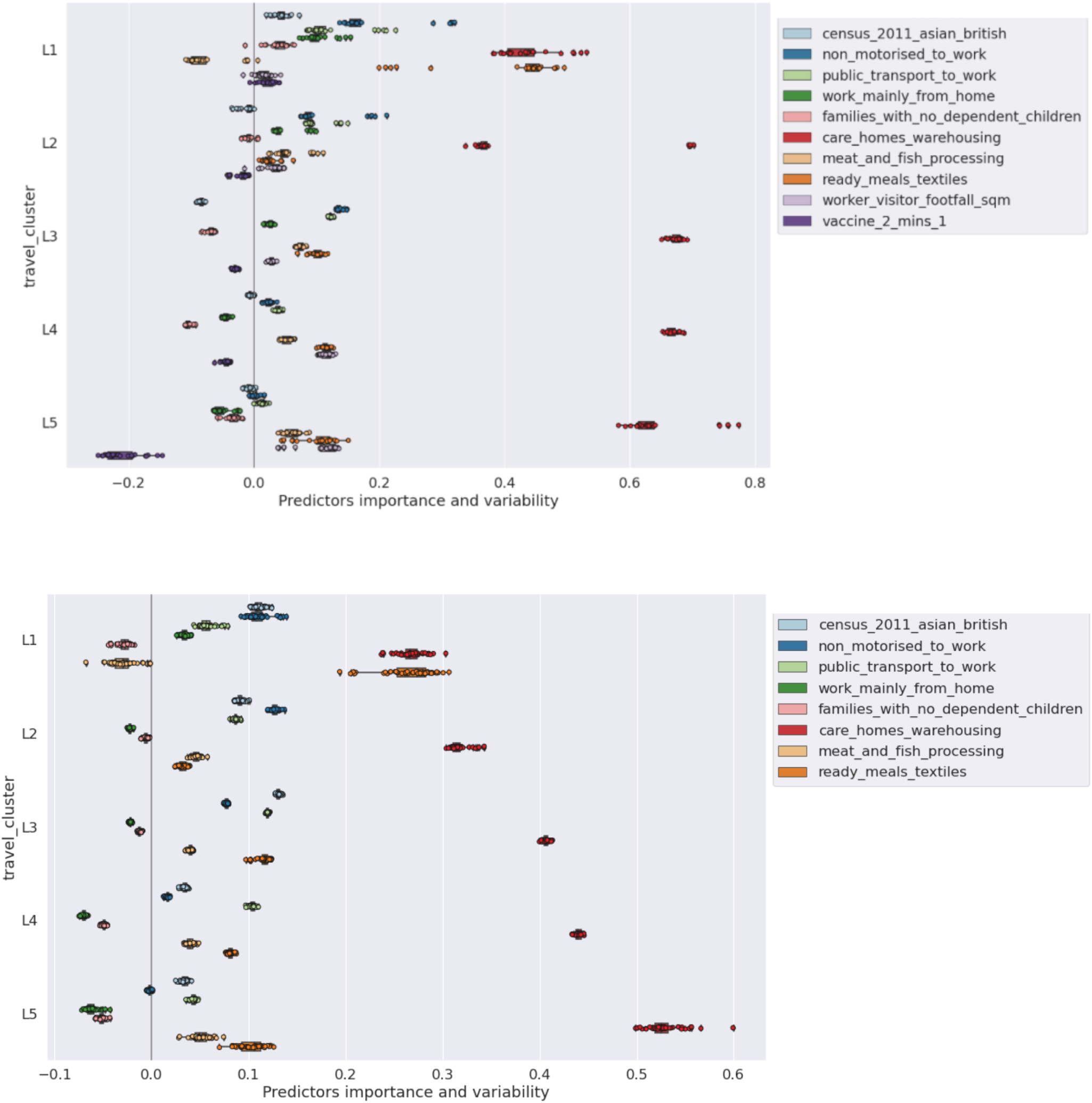
Variability of risk predictors used in our model. **(a)** Static and dynamic risk predictors variability for different travel clusters for the period 18^th^ July 2021-5^th^ December 2021 covering the period following the lifting of lockdown restrictions in England. **(b)** Static risk predictors variability for different travel clusters for the period 4^th^ October 2020-5^th^ December 2021 covering all time tranches.

Figure 5a shows the outcome of stability analysis of both the static and dynamic influences for the last time tranche 7. Tranche 7 covers the time period of the lifting of lockdown restrictions in England (increased mobility) and also when the cumulative coverage of the proportion of fully vaccinated population is highest across all travel clusters. The results suggest fairly stable and robust patterns of influences aligned with the findings presented in Figure 4.

Figure 5b shows the results of this cross validation exercise for the static risk influences across all time periods combined. It is interesting to note that static influences are relatively robust to temporal variations when modelled at geographically aggregate level and segmented by travel clusters. For instance, high-risk industries have stayed highly significant influence across all time periods modelled and travel clusters with positive impact on infection risk^11^. This suggests that policy interventions, although might have controlled the total level of infections across all communities, have not shown much influences on the relative risks specifically for the vulnerable communities working in high-risk industries.

In summary, we make the following observations from stability analysis at all modelled time periods:

- Large density of residents working in high-risk industries are a positive risk factor of infections in all land use settings.
- Larger density of smaller households with no dependent children are a negative risk factor for infections.
- Larger density of public transport and non-motorised users for commuting is a positive risk factor of infections for most dense urbanised areas.
- Residential areas with high density of those who tend to work from home are negatively associated with the risk of infections except for travel clusters L1 (Inner and Central London).

### 4.3 Model performance

We use R^2^ score to evaluate the performance of our model following the training on the static and dynamic variables for each tranche. The R^2^ score explains the dispersion of errors of a given dataset and can be used to measure the discrepancy between a model and actual data^23^. Scores close to 1.0 are highly desired, indicating better squares of standard deviations of errors. Figure 6 shows the R^2^ score for the fitted estimator for each time tranche of training and for every travel cluster. It is worth mentioning that the performance of our fitted estimators can be potentially improved by including additional variables in our modelling including the antibodies datasets which have not been available to us. It can be seen from Figure 6 that fitted estimators are better at capturing the spatial-temporal distribution of cases for less densely populated and rural areas (highest average R^2^ score).

**Figure 6.**
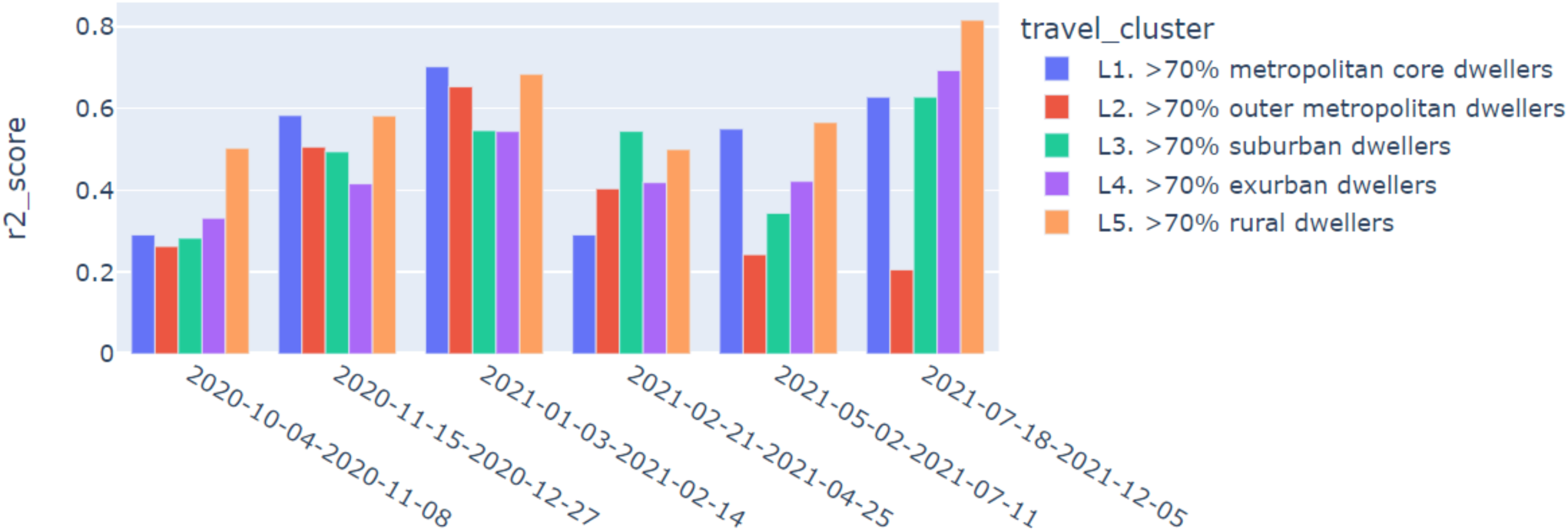
Goodness of fit captured through R^2^ score using the multi-group regression analysis for different travel clusters encapsulating different stages of the pandemic.

### 4.4 Limitations, mitigation, and future research plans

The analyses above provide some in-depth insights into the most important SARS-CoV-2 infection risk parameters at a granular spatial level and their spatiotemporal variations. To the best of our knowledge, this is the first study of its kind where a range of complementary datasets from various sources including vaccination and telecoms data have been used to understand the influences on SARS-CoV-2 infection risk.

Like all studies of this type, however, the research has many limitations, the potential effects of which are discussed in this section. We also discuss some of the potential avenues of further explorations towards expansion and implementation of the current work as a comprehensive framework of an early warning system at LSOA level for the UK.

One of the major limitations of our study is the potential ascertainment bias^27^ specifically in self-reporting datasets such as the test and trace. Our target variable can suffer from ascertainment bias despite wider testing available since the summer 2020. Recent studies^28^ have reported on the idea that using randomised testing schemes, such as the REACT study in the UK, can help debias fine-scale targeted testing data in order to provide accurate localised estimates of the number of infectious individuals.

Our modelling approach, however, could help accounting for some of these biases; using aggregated data means we focus on spatial (LSOA level) variations in cases rather than individual level information. This would make analysis less sensitive to biases arising from asymptomatic diseases which are likely to be underreported. In analysing spatial distribution of infection, one can assume high spatial correlation between asymptomatic and symptomatic infections making the spatial distribution of the latter a good representative of that for the former. Nonetheless, a detailed analysis to correct for potential biases in our input dataset can be an interesting avenue for future exploration.

A potential extension of the present investigation could be to expand the current analysis to include other occupation industries associated with different SARS-CoV-2 infections. The industries included in the present study were selected based on our literature review on infection rates by industry types and discussions we had with the UKHSA and the HSE. The extension of our current formalism to include other work sectors is relatively straightforward and is indeed considered by HSE as part of their work on monitoring workplace outbreaks.

Furthermore, to improve the model interpretability, which is the target of the current paper, we have adopted a multivariate linear regression and accounted for potential nonlinearity through further segmentation by travel clusters and time and consideration of interactive terms. The model predictability^12^, however, can be potentially improved (specifically after incorporation of further dynamic data discussed above) by introducing more advanced statistical models such as Structural Equation Model (SEM)^29^ to capture systematically interrelations between parameters (including conditional dependencies between cases, hospitalisation and mortality) and two-way fixed effects regression^30^, which can help causal inferences as well as short-term forecasting. Two-way fixed effect involves two modelling steps: a) modelling the prevalence of SARS-CoV-2 infection only on the static variables and b) estimating the impact of changes in dynamic variables on those in residuals from step (a) (i.e. the remaining effects after removing those from static variables). Estimating the residuals by dynamic variables can be incorporated under a linear or non-linear approximation including through use of machine learning techniques such as LSTM ^13^. This approach can facilitate short term and near real time risk inferences and forecasting.

### 4.5 Summary of key findings

The key findings of this work are highlighted below. First, our work has shed light on assessing the impact of the pandemic on some of the most vulnerable sections of the working population including those who work in high-risk industries, with relatively higher risk of exposure to infection. Our results, after controlling for real time mobility, vaccination and socioeconomic and demographic profiles, show that areas with a larger proportion of residents working in care homes and warehouses and to a lesser extent ready meals and textile sectors are prone to higher risk of infection across all travel clusters and all time periods. Similar influences across all residential area clusters suggests the potential association with workplace risk and regulations which can be further examined in a more detailed (individual level) workplace outbreak analysis.

Second, the findings underline the critical importance of geographical variations in influences on the prevalence of SARS-CoV-2 infection (after controlling for mobility and vaccination rate). For instance, for the most recent time tranche covering the period of lifting of lockdown restrictions in England, areas with a bigger proportion of small families and fewer children are prone to lower risk of infection. This is not a universal observation though and the respective risk is significant only in medium and smaller urban and rural areas but not in central and inner London and metropolitan cities. This is also the case for areas comprising of a larger proportion of those who can work from home; while work from home is shown to reduce the infection risk in less populated and smaller cities, this is not the case in metropolitan core dwellers where people might be more active in a diverse set of activities apart from work.

Finally, except rural settlements, areas with residents who are more dependent on the use of public transport for commuting have also been identified with greater risk of infections across all travel clusters. Although the risk is lower in the fifth tranche of time when vaccination has started to take effect and the Delta variant has not yet become dominant, use of public transport has been one of the main risk factors in most urban areas.

Given the above, our spatially aggregated model is well suited to tailor the research questions for further investigation at more detailed and granular level (e.g. through epidemiological models at individual and household levels). For instance, following our finding about the critical importance of certain industries’ workplace risk after controlling for the land use characteristics and mobility patterns at the residential area, further analysis can be tailored to design and evaluate safety measures and regulations in the future. Continuous evaluation of community level risk can identify new threats and risk patterns at an early stage when there is a better chance to respond.

### 4.6 Conclusion

In this work we have developed a relatively granular LSOA level SARS-CoV-2 infection risk model by bringing together a variety of datasets at LSOA level, in order to provide risk estimates of SARS-CoV-2 infections for neighbourhoods. Using LSOA level indicators encompassing population demographic, information on higher risk industries, housing conditions, urban/rural area classification, vaccination rates, and real time mobility patterns, we developed one of the most comprehensive dataset to date to model major community risk factors of SARS-CoV-2 infections in England. To fuse learning from this detailed dataset and control for exogeneities arising from the highly interrelated influences, we adopted machine learning and econometric techniques in our analysis pipeline on Google Cloud Platform. We used Latent Cluster Analysis (LCA) to identify distinct travel clusters within which travel patterns, behaviours and attitudes are more homogeneous. Training the model for each cluster separately, we applied multivariate regressions to gauge the static and dynamic influences and infer the major risk factors of the prevalence of SARS-CoV-2 infection. Our model can be updated regularly and run in real time to uncover the most recent risk factors driving the SARS-CoV-2 infections and associated variations in geographical patterns of risk. For the purpose of this paper, however, we used the model for seven distinct time periods which can best reflect the virus variants and policy interventions in the recent past. Our comprehensive identification of the risk factors affecting SARS-CoV-2 infections may be useful to policymakers to aid them devise effective non-pharmaceutical interventions besides medical ones for population groups most at risk and observe in real time the impact of global and federated policy interventions in mitigating the risk of SARS-CoV-2 infection.

## Data Availability

Some of the datasets used in this study are publicly available.

## Acknowledgment

We would like to thank Dr Thanasis Anthopoulos, for reviewing and quality assurance of the models’ code, Dr Arthur Turrell, Dr Louisa Nolan, and Dr Owen Daniel, for reviewing the manuscript and their very constructive and comprehensive comments. We also like to thank Alistair Calder and Chris Gale from the ONS Geospatial Analysis team for their support in data preparation of a wide range of static indicators tested or used in the model. Alongside, we want to thank colleagues from the Health and Pandemic Insights Team at ONS for their feedback and suggestions on the manuscript. We are also grateful to the colleagues in HSE and UKHSA for their feedback, constructive discussions and help in data acquisition over the course of this investigation.

## A Appendix A: variable definitions and data sources

Variable definitions, data dictionary and list of variables used in the LSOA level risk model.

**Table A.1.** A hierarchy of ONS census geography boundaries for the UK. Please refer to https://www.ons.gov.uk/methodology/geography/ukgeographies/censusgeography for more information on definitions of CENSUS geography including LSOA.

| Geography | Count (England & Wales) | Hierarchy |
| --- | --- | --- |
| Output Area (OA) | 181,408 | within LSOA |
| Lower Layer Super Output Area (LSOA) | 34,753 | within MSOA |
| Middle Layer Super Output Area (MSOA) | 7,201 | within Local Authority (LA) |
| Workplace Zone (WPZ) | 53,578 | constrained to MSOA |

**Table A.2.**
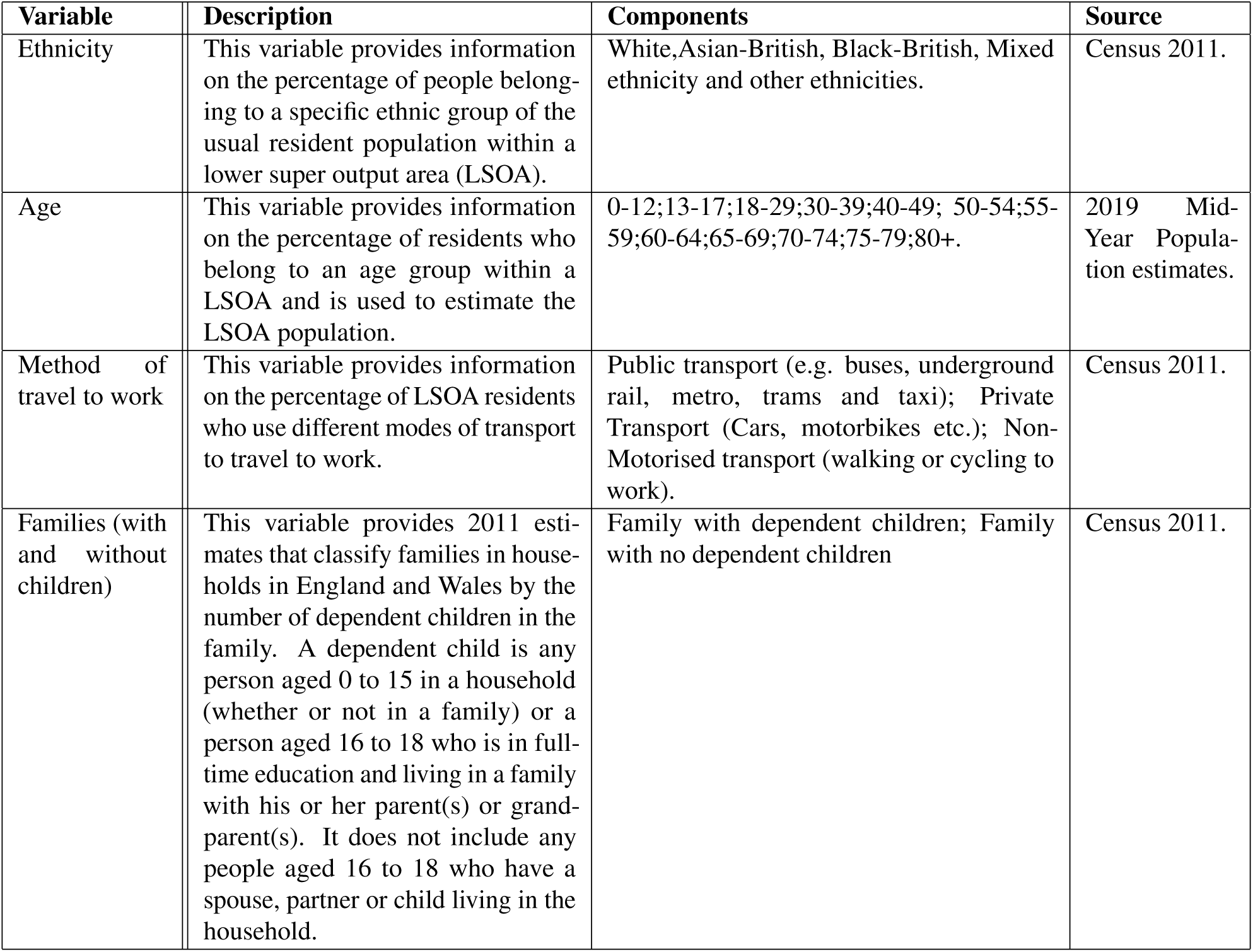
Data dictionary outlining all the static variables used in the LSOA level risk model and the respective data sources.

**Table A.3.** List of variables and associated datasets used in the LSOA level risk model.

|  |
| --- |
| Static and dynamic datasets used in the model |

| Input variable at residential end | Datasets used/ published department | Geography<sup>14</sup> | Temporal |
| --- | --- | --- | --- |
| Travel Clusters extracted from our earlier work<sup>11</sup> | Individual level National Travel Survey (NTS)- 2002 to 2015 / DfT | LSOA | Static |
| Ethnicity, Method of travel to work, Family structure (with and without children), Journey to work (JTW) | Census 2011, 2019 Mid-Year Population Estimate / ONS | LSOA | Static |
| High risk industries' workers | Inter-department Business Registry (IDBR) 2019 / ONS | LSOA | Static |
| Workers and Visitors Footfall | Deimos footfall dataset / Telefonica (O<sub>2</sub>) | MSOA (dis-aggregated to LSOA using worker proportion as weights e.g. if 20% of MSOA's workers work in an LSOA, 20% of the MSOA footfall will be allocated to that LSOA) | Dynamic |
| Vaccination uptake-first and second doses administered | National Immunisation Management Service (NIMS) vaccination dataset / NHSD | LSOA | Dynamic |
| confirmed COVID-19 positive cases (Y Target variable) | Second Generation Surveillance System (SGSS) positive tests dataset / PHE. | LSOA | Dynamic |

**Table A.4.** SIC codes for a list of high-risk industries.

| High risk industry | SIC CODE | SIC NAME |
| --- | --- | --- |
| Meat and fish processing | 10.1 | Processing and preserving of meat and production of meat products |
| Meat and fish processing | 10.2 | Processing and preserving of fish, crustaceans and molluscs |
| Textiles | 13.1 | Preparation and spinning of textile fibres |
| Textiles | 13.2 | Weaving of textiles |
| Textiles | 13.3 | Finishing of textiles |
| Textiles | 13.9 | Manufacture of other textiles |
| Textiles | 14.1 | Manufacture of wearing apparel, except fur apparel |
| Textiles | 14.2 | Manufacture of articles of fur |
| Textiles | 14.3 | Manufacture of knitted and crocheted apparel |
| Textiles | 15.1 | Tanning and dressing of leather; manufacture of luggage, handbags, saddlery and harness; dressing and dyeing of fur |
| Textiles | 15.2 | Manufacture of footwear |
| Care | 87.1 | Residential nursing care activities |
| Care | 87.2 | Residential care activities for learning disabilities, mental health and substance abuse |
| Care | 87.3 | Residential care activities for the elderly and disabled |
| Care | 87.9 | Other residential care activities |
| Warehousing | 52.1 | Warehousing and storage |
| Ready meals | 10.85 | Manufacture of prepared meals and dishes |

**Table A.5.** Distinct time tranches alongside the main external influence dominating a specific tranche.

| Tranche | Period | External influences during different stages of the pandemic |
| --- | --- | --- |

<sup>14</sup>Geography entails ONS census geography boundaries for the UK. More information can be found here: https://www.ons.gov.uk/methodology/geography/ukgeographies/censusgeography

|  |  |  |
| --- | --- | --- |
| 1 | 2020-05-03 to 2020-08-30 | Low prevalence; schools closed; Alpha and Delta variants not yet emerged; no vaccine available. |
| 2 | 2020-09-06 to 2020-11-08 | High prevalence; schools open; negligible Alpha variant; Delta variant not yet emerged; no vaccine available. |
| 3 | 2020-11-15 to 2020-12-27 | High prevalence; schools open; Alpha variant becomes dominant; Delta variant not yet emerged; negligible vaccine coverage. |
| 4 | 2021-01-03 to 2021-02-14 | High prevalence; schools closed (except for pre-school); Alpha variant dominant; Delta variant not emerged yet; over 10 million first vaccine doses by the end of the time period. |
| 5 | 2021-02-21 to 2021-04-25 | Low prevalence; schools open; Delta variant negligible; over 35 million first and 15 million second vaccine doses by the end of time period. |
| 6 | 2021-05-02 to 2021-07-11 | High prevalence; schools open; Delta variant becomes dominant; over 45 million first and 35 million second doses administered by the end of the time period. |
| 7 | 2021-07-18 to 2021-12-05 | Lifting of almost all lockdown restrictions in England and before the Omicron variant became dominant. |

## B Appendix B: Descriptive analysis

In this Appendix we highlight the findings from exploratory factor analysis (EFA) for static predictors; this helps group the input variables where they are highly correlated to account for their spatial interactions; second, we present the distribution of input variables across the five distinct travel clusters; this further affirm the use of multi-group approach (i.e. fitting separate model for each cluster) to account for spatial sorting and self-selection effects.

### B.1 Exploratory factor analysis and spatial interactions of input variables

Table B.1 shows the factor loadings from EFA; we have only reported those with significant size-i.e. an absolute loading cut-off of 0.35. Factors in essence represent common communality across variables and the associated factor loadings (which are ranged from −1 to 1) show the extent to which each variable has contributed to each factor. This means the variables with large absolute factor loadings for each factor tend to correlate with each other and the associated factor can represent their interrelations. For instance, in our analysis below, we can see relatively large factor loadings for the first factor for black, mixed and other ethnic groups and for commuting with public transport (all above 0.7). This shows strong spatial correlation amongst these variables suggesting higher probability for these ethnic groups to live in places with relatively good public transport access for commuting (i.e. mainly London and metropolitan areas). A complete list of spatially correlated variables inferred for each individual Factor is shown in Table B.2.

**Table B.1.** Results of exploratory factor analysis of the static variables where ∗∗ is used to indicate the respective factor loading is below the chosen threshold of 0.35.

| variables | Factor_1 | Factor_2 | Factor_3 | Factor_4 |
| --- | --- | --- | --- | --- |
| Census 2011 Asian British | 0.45 | ** | ** | 0.46 |
| Non-motorised to work | ** | ** | ** | ** |
| Public transport to work | 0.85 | ** | ** | ** |
| Work mainly from home | ** | -0.35 | -0.49 | ** |
| Families with no dependent children | -0.39 | ** | -0.72 | ** |
| Care homes | ** | 0.69 | ** | ** |
| Meat and fish processing | ** | ** | ** | ** |
| Ready meals | ** | ** | ** | 0.42 |
| Textiles | ** | ** | ** | 0.50 |
| Warehousing | ** | 0.40 | ** | ** |
| Census 2011 Mixed multiple ethnicity | 0.84 | ** | ** | ** |
| Census 2011 other ethnicity | 0.71 | ** | ** | ** |
| Census 2011 Black-African, Caribbean, Black-British | 0.78 | ** | ** | ** |

**Table B.2.** Descriptions of the different Factors obtained after EFA.

| Factors | Description |
| --- | --- |
| Factor_1 | Strong spatial correlation amongst populations groups from Black (African, Caribbean and Black British), Mixed and Other ethnicities. Alongside strong (spatial) correlation with regions where the use of public transport to work by the residents is higher. |
| Factor_2 | Spatial correlation amongst residents working in care homes, and warehouses. |
| Factor_3 | Spatial correlation amongst regions with larger concentration of families with no children and larger proportion of the population working from home. |
| Factor_4 | Spatial correlation amongst residents working in textiles and ready meals. |

Based on the above inference from EFA as reported in Table B.2, we make the following assumptions for our modelling stage:

- We use the proportion of the working population using public transport as the representative variable for Factor_1 and drop other highly correlated variables comprising Factor_1 from our variable list. This is to avoid multicollinearity issues.
- We define a new variable capturing the total resident population working in care homes, and warehousing industries.
- We define a new variable capturing the total resident population working in ready meals and textiles industries.
- We incorporate residents working in meat and fish processing as independent variables in our analysis.
- For each LSOA, we also include the proportions of families with no children, population working from home, population using non-motorised transport to work and British-Asian ethnic groups as independent predictors in our analysis.

The alternative approach would have been incorporating latent factors into our model instead of choosing one of the correlated variables or combine those with each other; however, the latter that we have adopted made our findings more interpretable for the purpose of this paper.

**Figure B.1.**
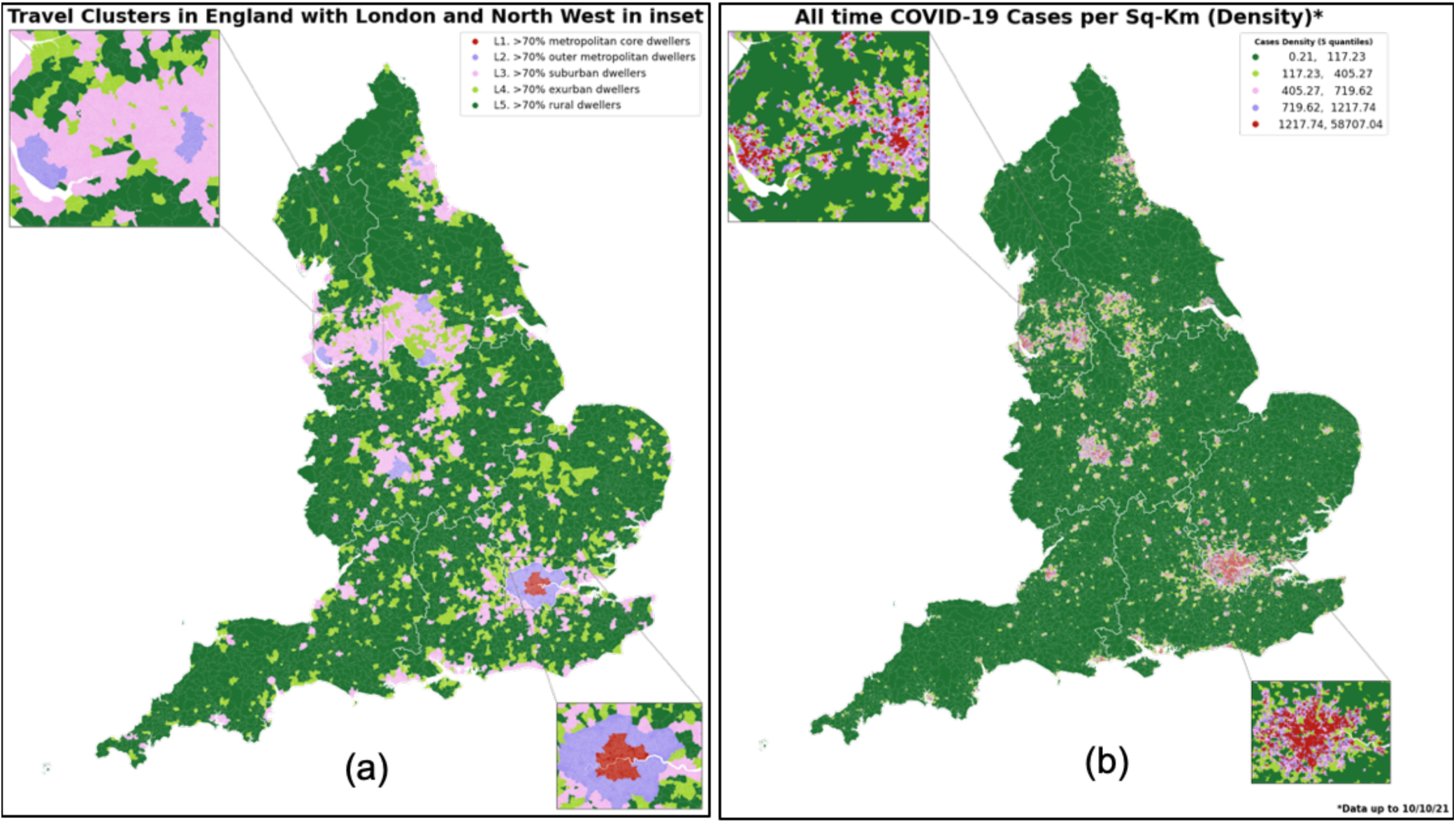
(a) Spatial distribution of latent travel clusters (b) spatial distribution of density of COVID-19 cases over the period of the analysis.

**Figure B.2.**
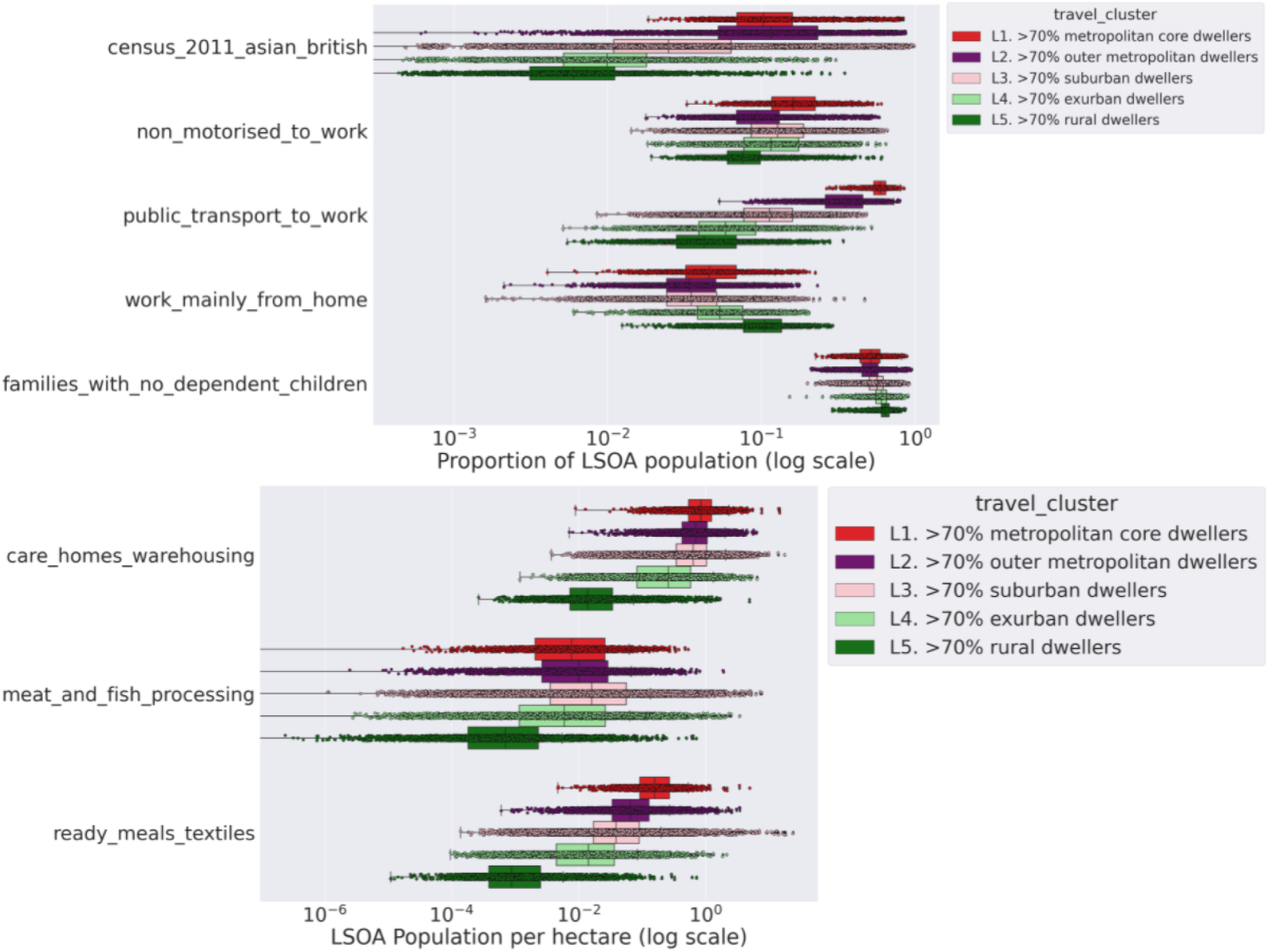
Distribution of the static predictors used in the analysis across distinct travel clusters.

### B.2 Descriptive analysis of spatial distribution of input variables

In this section, we present the geographical distribution of input variables across the five identified travel clusters. Figure B.1 compares the geographical distribution of travel clusters (Figure B.1 (a)) with that of COVID-19 cases (Figure B.1 (b)) over the period of analysis (i.e. from 4^th^ October 2020-5^th^ December 2021). It can be observed that travel clusters are already reflecting geographic variations in COVID-19 cases in many areas, although there are yet important differences as well.

Figure B.2 and Figure B.3 show the distribution across spatial clusters of the static and dynamic variables and suggests clear distinctions in socioeconomic and demographic as well as mobility patterns across travel clusters. This shows not only can travel clusters capture some of the important spatial interactions but also it can help account for spatial sorting and self-selection of socioeconomic and demographic groups. In other words, through segmenting by travel clusters, our multigroup analysis allow comparing like with like.

It is worth highlighting that there exists spatial correlation amongst residents working in care homes, and warehouses and this is why we have defined a new single variable capturing the total resident population working in care homes and warehousing industries as shown in Figure B.2. Same is true for the number of LSOA residents working in ready meals and textiles industries. For further details refer to results presented in Section B.1.

In particular, we can infer the following from Figure B.2:

- More dense urbanised areas have the highest proportion of people from an Asian background
- Use of public transport and non-motorised transport to work is also highest amongst the dense urbanised areas.
- Proportion of the working population more likely to work from home is highest amongst more rural areas.

**Figure B.3.**
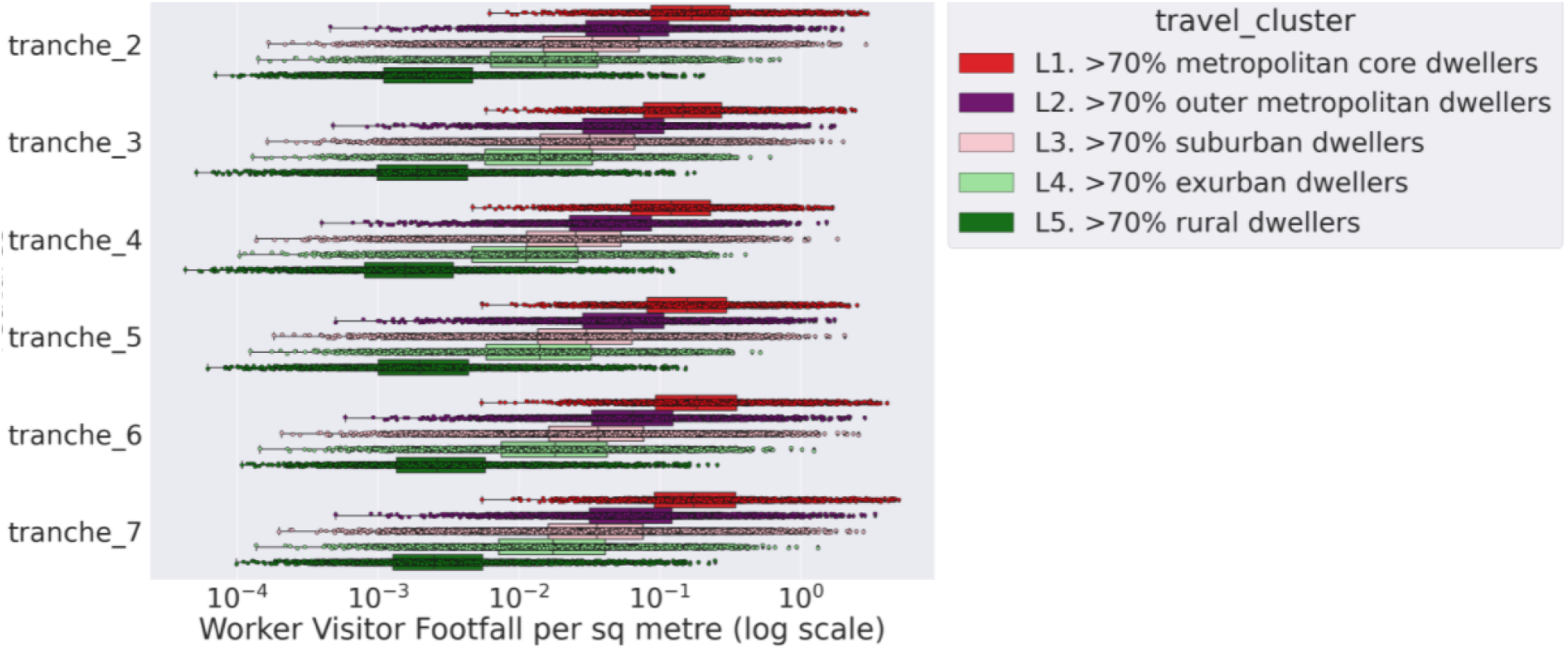
Distribution of the worker and visitor footfall across distinct travel clusters for distinct time tranches studied under this investigation **. ** Note: the data sharing agreement with our Mobility footfall data provider does not include the period under tranche 1.
- Proportion of households with no children is comparable across urban and rural areas.
- Resident population working in high-risk industries is highest amongst the most densely populated urban areas except for workers in meat and fish processing who have slightly higher tendency to live in medium and smaller urban areas but not much in rural areas.

With respect to dynamic variables, Figure B.3 shows that more dense urbanised areas also have the highest average footfall of workers and visitors for each distinct time tranche (refer to Table A.5 for time tranche definitions). In addition, Figure B.4 shows the cumulative proportion of population administered with first and second doses of COVID-19 vaccinations for each travel cluster and successive time tranches ^15^; this suggests that vaccination uptake and rates is higher in more rural areas compared to more dense urbanised areas. This probably is due to the difference in age profiles of residents in urban and rural areas with the latter tend to have older population who got priority in the vaccination roll out.

**Figure B.4.**
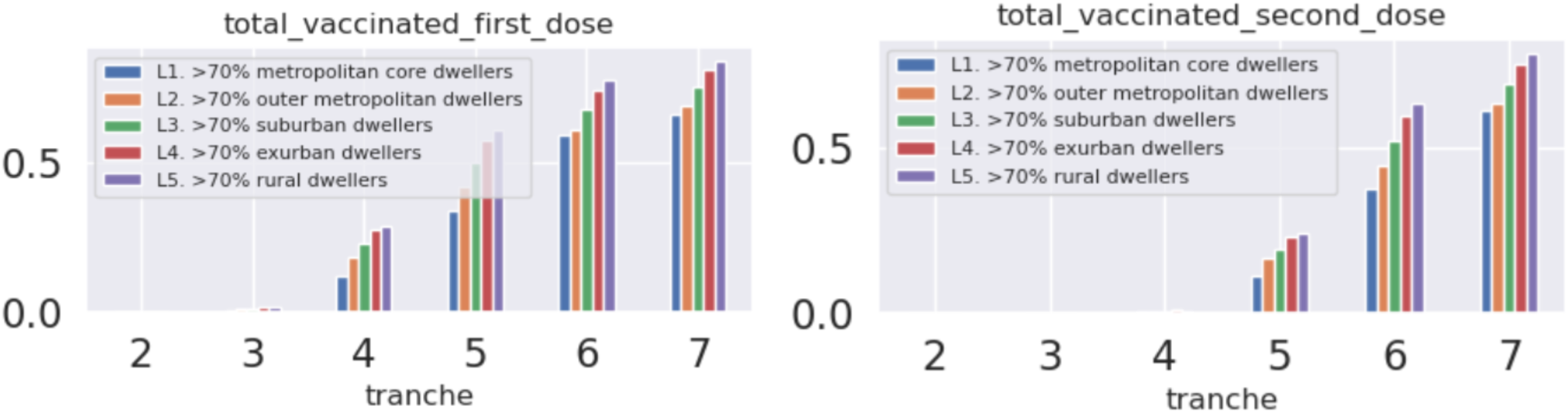
Proportion of population administered with first and second doses of COVID-19 vaccinations for each travel cluster.

**Table C.1.**
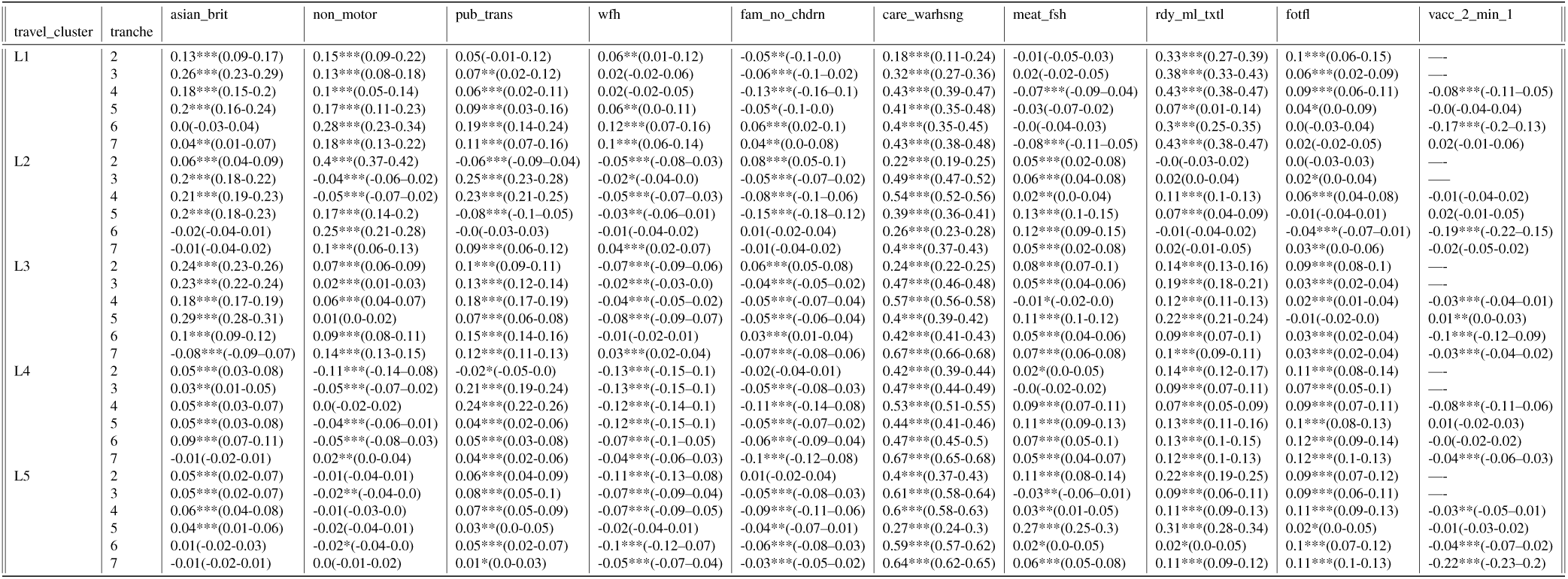
Standardised risk predictors influencing infections in England covering the period 4^th^ October 2020-5^th^ December 2021 *∗∗∗p <* 0.01*, ∗∗ p <* 0.05*, ∗p <* 0.1. The predictors have been explicitly defined in Table 2. Also provided are the 95% confidence interval for each coefficient.

## C Appendix C: standardised risk predictors

Table C.1 shows the standardised coefficients across all time tranches. Please note that one should compare coefficients at each row as they are standardised for each travel cluster and time tranche to help compare relative importance of each influences within every travel cluster and time tranche.

## D Appendix D: non-standardised risk predictors

This appendix provides variations in influences over time across travel clusters; as the risk influences are expressed as non-standardised coefficients (with units), influences within each travel cluster are not comparable against each other. However, the evolution of a risk influence over time within each travel cluster can still be inferred as shown in Figure D.1.

**Figure D.1.**
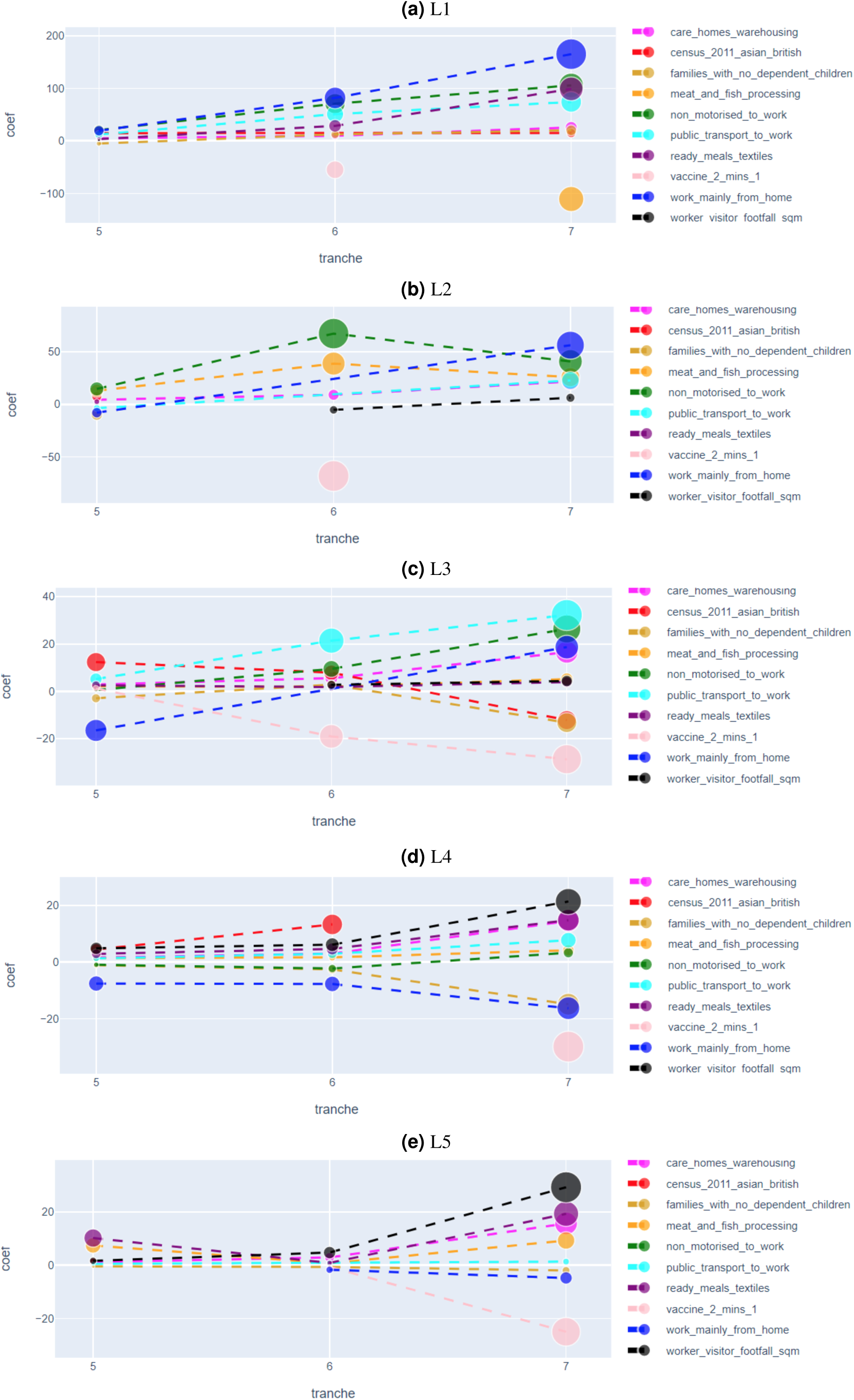
Evolution of standardised regression coefficients for static and dynamic predictors over the various tranches of interest.

## E Appendix E: Prediction of Risk for an early warning detection framework

In the previous section, we aggregated the static and dynamic predictors and inferred the most significant risk factors driving infections for each individual travel cluster. To further leverage the usefulness of the model, we use the trained estimator to predict the future number of cases on unseen test dataset.

For this we use a “sliding window” training approach where we can make use of an estimator fitted on a prior time tranche to predict the number of cases for the next time tranche. It is worth pointing out that the size of the time-window of the test tranche is governed by the time span of the dynamic predictors alone, and can be somewhat arbitrary. The test tranche can span over a single week or longer if desired. If the test dataset covers a time period significantly different from the train data, the predictions from the model are expected to be different from the actual number of cases. For example, the Omicron variant was not dominant in England for the period covered under tranche 7, and estimator trained on the dataset for tranche 7 is expected to perform poorly in terms of predictions for the successive tranche covering the period of enhanced transmissions driven by the Omicron variant. Deviation from the models’ predictions can be an important indicator in flagging areas of potential interest. For example, LSOAs with significant positive deviation (underestimation by the model) can be brought to the attention of decision makers as emerging hotspots where the growth of the infection curve cannot be accounted for despite controlling for a range of static and dynamic predictors investigated in this study.

To illustrate the above idea further, we fit an estimator for each travel cluster using the dynamic and static predictors aggregated between the period 18^th^ July 2021-5^th^ December 2021 (tranche 7). Following the training, we use the estimators to predict future cases per unit area. We also apply regularisation to tackle the issue of potential overfitting and variable selection in the training step. The predicted number of cases per unit area are shown in a thematic map in Figure E.1. As evident, the highest number of infections (per unit area) are expected to occur in the most dense urbanised area. Of the top 20 LSOAs with the highest predicted number of cases, the majority of the LSOAs are in London signalling some of the highest infection rates prevalent in most dense urbanised areas.

**Figure E.1.**
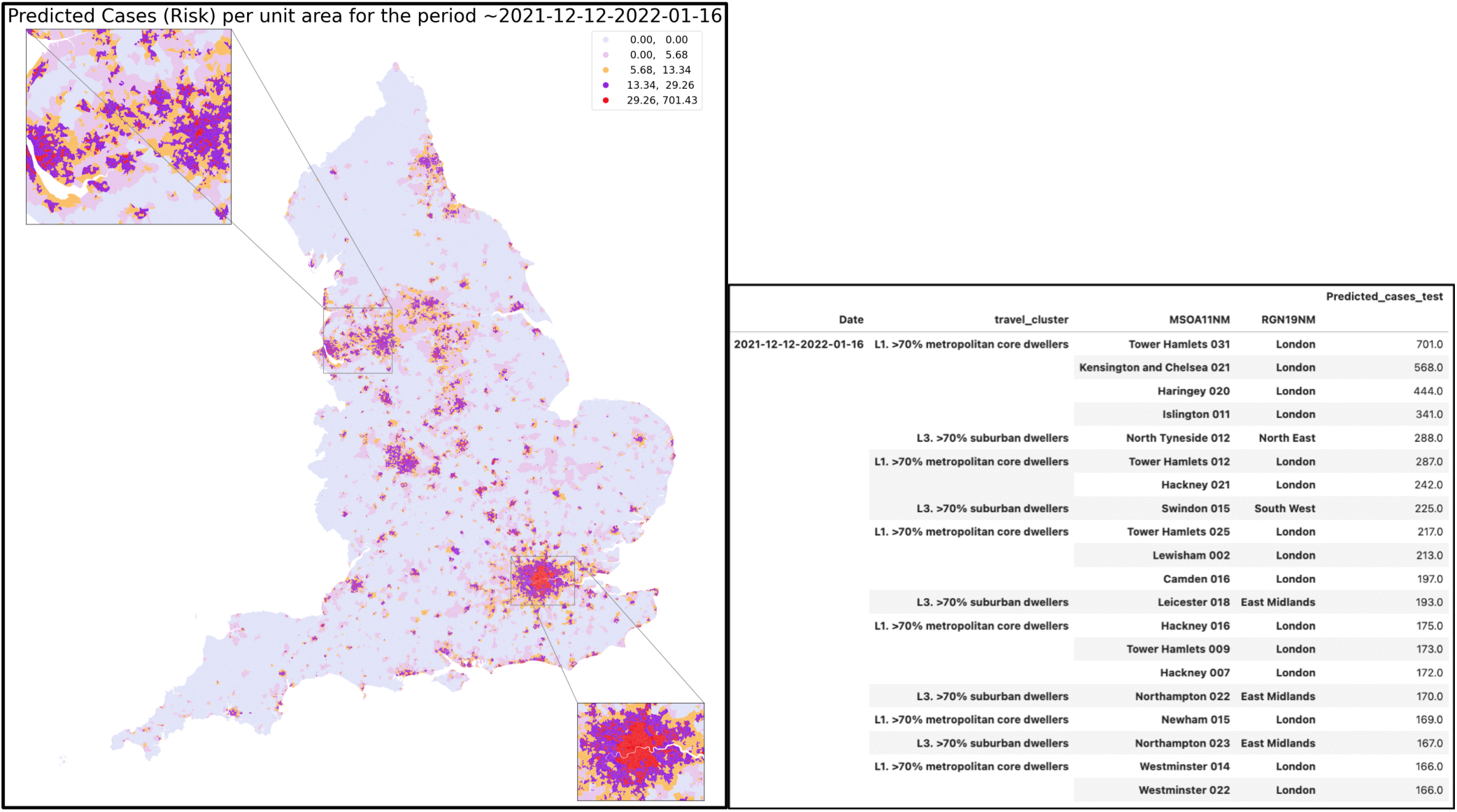
Predicted mean number of cases per unit area on the latest test data encompassing the period 12^th^ December 2021-16^th^ January 2022. Also shown are the top-20 LSOAs with the highest predicted cases per unit area for the corresponding time period ^*-*^. ^*-*^ The model was trained on the dynamic and static predictors aggregated between the period 18^th^ July 2021-5^th^ December 2021. The training dataset covers the period of no national level restrictions in England except the introduction of facial covering in certain public places and on public transport wef 30^th^ November 2021. The Omicron variant was dominant in the period covered by the test data.

## Footnotes

1 A hierarchy of ONS census geography boundaries for the UK is presented in Table A.1 in Appendix A.

2 The distribution of socioeconomic patterns and land use features such as population density or accessibility to amenities tend to be relatively stable. It can take decades for changes in population profile (such as tendency to live in less populated area for younger population due to increased appetite to work from home) to take place, therefore, Census 2011 can still capture well the land use patterns and geographical distribution of population by their socioeconomic and demographic patterns.

3 Workers who carry out work consisting of residential nursing activities, Residential care activities for learning disabilities, mental health and substance abuse, residential care activities for the elderly and disabled, and other residential care activities

4 We found that Leicester (which was a COVID-19 hotspot at that time) had one of the highest population density of workers working in the chosen work sectors.

5 Workplace Zones are an output geography, produced using workplace data from the 2011 Census for England and Wales. They are designed to supplement the Output Area (OA) geographies which were introduced with the 2001 Census, and have been constructed from OAs, or sub-divisions of these called postcode-level building-blocks (PCBBs). While OAs are designed to contain consistent numbers of persons based on where they live, WPZs are designed to contain consistent numbers of workers, based on where people work. The dataset can be downloaded from the census official website (https://www.nomisweb.co.uk/output/census/2011/wf02ew_oa.zip).

6 Under pillar 2 testing route, swab testing is conducted for the wider population as set out in the UK government guidance.

7 PHE has confirmed that data included home tests through lateral flow devices (LFDs) alongside lab results.

8 The comprehensive list of socioeconomic and demographic variables used from the NTS data are provided in Jahanshahi and Ying, 2021^11^

9 One can apply this model for regular and real time prediction of infections at area based level which was one of the tasks at hand when this work was originally commissioned at the height of the COVID-19 pandemic. This, however, is not reported in this paper which has a focus on reporting inference from analysis.

10 Since the dynamic influences (vaccination and footfall) tend to vary over time, the analysis of the stability of coefficients over all time tranches is done only for static variables.

11 The exception is the influence of meat and fish processing in Central and Inner London cluster which can be explained by the small sample size of workers in meat and fish processing in Central London.

12 See Appendix E for a brief analysis on how our current formalism can be potentially used as an early warning detection framework for risk mitigation.

13 Long short-term memory

14 Geography entails ONS census geography boundaries for the UK. More information can be found here: https://www.ons.gov.uk/methodology/geography/ukgeographies/censusgeography

15 The source of vaccination data (NIMS) and population at LSOA (MYE) are different and hence the proportion of those vaccinated (vaccination normalised by population) we used for this paper might be subject to some level of biases. However, we believe that the spatial distribution of those across LSOAs (relative values), which is the major input to our model, should be more robust and suitable for our analysis. We also use the changes from first dose uptake to the second as our input variable which can further help cancelling out some of the potential inherent biases in data.

## References

1. Steel, K. & Yapp, R. Office for national statistics. coronavirus (covid-19) infection survey: methods and further informa-tion. https://www.ons.gov.uk/peoplepopulationandcommunity/healthandsocialcare/conditionsanddiseases/methodologies/covid19infectionsurveypilotmethodsandfurtherinformation (2022).

2. House, T. & et. al. Inferring risks of coronavirus transmission from community household data. arXiv e-prints:2104.04605 DOI: https://arxiv.org/abs/2104.04605 (2021).

3. Williamson, E. J. & et. al. Factors associated with covid-19-related death using opensafely. Nature 584, 430–436, DOI: 10.1038/s41586-020-2521-4 (2020).

4. Katikireddi, S. V. & et. al. Unequal impact of the covid-19 crisis on minority ethnic groups: a framework for understanding and addressing inequalities. J Epidemiol Community Heal. 75, 970–974, DOI: 10.1136/jech-2020-216061 (2021).

5. Raisi-Estabragh, Z. & et. al. Greater risk of severe covid-19 in black, asian and minority ethnic populations is not explained by cardiometabolic, socioeconomic or behavioural factors, or by 25 (oh)-vitamin d status: study of 1326 cases from the uk biobank. J. Public Heal. 42, 451–460, DOI: 10.1093/pubmed/fdaa095 (2020).

6. Jin, J. & et. al. Individual and community-level risk for covid-19 mortality in the united states. Nat. medicine 27, 264–269, DOI: 10.1038/s41591-020-01191-8 (2021).

7. Sze, S. & et. al. Ethnicity and clinical outcomes in covid-19: a systematic review and meta-analysis. Nat. medicine 29, 100630, DOI: 10.1016/j.eclinm.2020.100630 (2020).

8. Khunti, K. & et. al. Is ethnicity linked to incidence or outcomes of covid-19? BMJ 369, DOI: 10.1136/bmj. m1548 (2020).

9. Wang, X. Mapping and identifying community risk factors for covid-19 nursing home deaths. Heal. Serv. Res. 56, 85–85, DOI: 10.1111/1475-6773.13841 (2021).

10. Yapp, R. & et. al. Office for national statistics. coronavirus (covid-19) infection survey technical article: analysis of populations in the uk by risk of testing positive for covid-19. https://www.ons.gov.uk/peoplepopulationandcommunity/healthandsocialcare/conditionsanddiseases/articles/coronaviruscovid19infectionsurveytechnicalarticle/analysisofpopulationsintheukbyriskoftestingpositiveforcovid19september2021# (2021).

11. Jahanshahi, K. & Jin, Y. Identification and mapping of spatial variations in travel choices through combining structural equation modelling and latent class analysis: findings for great britain. Transportation 48, 1329–1359, DOI: https://link.springer.com/article/10.1007/s11116-020-10098-9 (2021).

12. Office for national statistics. open geography portal. https://geoportal.statistics.gov.uk/.

13. English indices of deprivation, department for levelling up, housing and communities and ministry of housing, communities & local government. https://www.gov.uk/government/statistics/english-indices-of-deprivation-2019 (2019).

14. Full ons 2011 census data for different categories can be found on the ons website. https://www.ons.gov.uk/.

15. Office for national statistics. inter-departmental business register (idbr). https://www.ons.gov.uk/aboutus/whatwedo/paidservices/interdepartmentalbusinessregisteridbr.

16. Kirk, A. & et. al. Data reveals coronavirus hotspots in bradford, barnsley and rochdale. https://www.theguardian.com/world/2020/jul/01/data-reveals-coronavirus-hotspots-in-bradford-barnsley-and-rochdale (2013).

17. Office for national statistics. classification of workplace zones for the uk methodology and variables. https://www.ons.gov.uk/methodology/geography/geographicalproducts/areaclassifications/2011workplacebasedareaclassification/classificationofworkplacezonesfortheukmethodologyandvariables (2011).

18. Office for national statistics. uk sic 2007. https://www.ons.gov.uk/methodology/classificationsandstandards/ukstandardindustrialclassificationofeconomicactivities/uksic2007 (2022).

19. Uk health security agency. covid-19 vaccine surveillance report. https://assets.publishing.service.gov.uk/government/uploads/system/uploads/attachment_data/file/1063023/Vaccine-surveillance-report-week-12.pdf (2022).

20. Kozlov, M. Waning covid-19 super-immunity raises questions about omicron. Nature 48, 1329–1359, DOI: https://www.nature.com/articles/d41586-021-03674-1 (2021).

21. Goldberg, Y. & et. al. Protection and waning of natural and hybrid covid-19 immunity. MedRxiv 48, 1329–1359, DOI: 10.1101/2021.12.04.21267114 (2021).

22. Pagel, C. & et. al. Tackling the pandemic with (biased) data. Science 374, 403–404, DOI: https://www.science.org/doi/10.1126/science.abi6602 (2021).

23. Johnson, R. & Wichern, D. Applied Multivariate Statistical Analysis (6th ed) (Pearson, Ch. 9, 2007).

24. More details of the package can be found here. https://factor-analyzer.readthedocs.io/en/latest/index.html.

25. What thresholds should i use for factor loading cut-offs? https://imaging.mrc-cbu.cam.ac.uk/statswiki/FAQ/thresholds.

26. Geron, A. Hands-on machine learning with Scikit-Learn, Keras, and TensorFlow: Concepts, tools, and techniques to build intelligent systems (OŔeilly Media, 2019).

27. E.J., G. & et. al. Estimation of ascertainment bias and its effect on power in clinical trials with time-to-event outcomes. Stat Med. 40, 1306–1320, DOI: 10.1002/sim.8842 (2021).

28. G., N. & et. al. Local prevalence of transmissible sars-cov-2 infection: an integrative causal model for debiasing fine-scale targeted testing data. medRxiv DOI: 10.1101/2021.05.17.21256818 (2021).

29. Kline, R. Principles and Practice of Structural Equation Modelling (2nd Edition ed.) (New York: The Guilford Press, 2005).

30. Imai, K. & Kim, I. On the use of two-way fixed effects regression models for causal inference with panel data. Polit. Analysis 29, 405–415, DOI: 10.1017/pan.2020.33 (2021).

